# Cost-Utility and Cost-Effectiveness Analysis of disease-modifying drugs of Relapsing-Remitting Multiple Sclerosis: A Systematic Review

**DOI:** 10.1101/2022.08.10.22278518

**Authors:** Nasrin Abulhasanbeigi gallehzan, Majid Khosravi, Samira Soleimanpour, Saeed Hoseini, Habibeh Mir, Vahid Alipour, Aziz Rezapour

## Abstract

**Background:** Multiple sclerosis is a chronic demyelinating disorder of the central nervous system that is categorized as an immune-mediated inflammatory disease. This study aimed to systematically review the cost-benefit and cost-effectiveness of relapsing-remitting drugs for multiple sclerosis.

**Methods:** To find related research and articles, articles published in Iranian and international databases by using a combination of MeSH (Medical Subject Headings) terms and based on inclusion and exclusion criteria were searched and reviewed. We included studies that addressed interventions, ICER, per QALY, and those which were published in a journal for different methods of infertility treatment, or a major general journal till 2019.

**Results:** Out of 1,360 records found, finally, 21 records were included in the research. Ten articles were published in the European continent, six articles in the Americ continent, and finally, five articles in the Asia continent. The most common limitations of published economic evaluation studies were in methodology or presentation of incremental analyses, sensitivity analyses, and discounts. The lowest and highest numerical value of outcome measures were -1,623,918 and 2,297,141.53, resprectively. Furthermore, the lowest and highest numerical value of the cost of disease-modifying drugs of RRMS were $180.67, and $1474840.19, resprectively.

**Conclusions:** Based on the results of all studies, it can be concluded that for the treatment of patients with AF, care-oriented strategies should be preferred to drug strategies. Also, among the drug strategies with different prescribing methods, oral disease-modifying drugs of RRMS should be preferred to injectable drugs and intravenous infusions.

## Introduction

Multiple sclerosis is a chronic demyelinating disorder of the central nervous system that is categorized as an immune-mediated inflammatory disease. (1, 2) The clinical course and severity of the disease are variable, however, in general, the common symptoms of the disease include paralysis, tingling, weakness, impaired balance and gait, blurred vision or diplopia, vertigo, cognitive impairment, fatigue, and urinary bladder dysfunction. (3) This disease consists of four stages, including recurrent multiple sclerosis, primary progressive multiple sclerosis, secondary progressive multiple sclerosis, and progressive multiple sclerosis. (4) The course of relapsing-remitting multiple sclerosis is characterized by worsening of neural functions, with the episode of unpredictable attacks, followed by periods of relative recovery. (5)

About 85% of people with MS are initially diagnosed with relapsing-remitting, which is characterized by destructive attacks that are quite noticeable on nerve function, followed by periods of complete or partial recovery and no progression of the disease. Over time, most people are diagnosed with a transition from relapsing-remitting period to secondary progressive multiple sclerosis. This transmission is manifested by the continuous progression of the disease without periods of recovery.

(6) As far as the incidence of this disease is concerned, by gender, women are two or three times more affected by this disease than men. (4) About 573,000 individuals in the United States are affected by multiple sclerosis. Most people with the disease are diagnosed at 20-50 years of age. (7)

The prevalence of MS in some parts of the world is high, for example, in North America at 140 per 100,000 and in Europe at 108 per 100 million. (4) Whereas the prevalence of multiple sclerosis in Thailand, reported by Prayon Vivat et al., was 201 per 100,000 people. (5) Lost productivity due to progressive disabilities contributes to the heavy economic burden of the disease. (8) It is important to note that the necessary medical care for MS patients and the significant cost of the pharmacy place a heavy economic burden on policymakers and caregivers. On average, patients with MS see their doctor eight times a year, which is about three times as many people without the disease. (9) Most studies have reported the costs of disease-modifying drugs (DMDs) to account for a large proportion of total medical costs (64% to 91%) in the treatment of MS. (10) The main goal of different MS treatments is to prevent or delay long-term disabilities. There is no final cure for MS. DMTs used to correct the course of MS and reduce or slow the progression of the disease were approved by the Food and Drug Administration (FDA) on Dec. 31, 2014, for the treatment of recurrent types of the disease. (6) Before 2010, injectable interferon beta and glatiramer acetate were the only treatment options available for patients with relapsing-remitting multiple sclerosis. These two drugs reduce recurrence by about a third, But they are less effective in slowing down the inability to face new factors. (11) These drugs are reimbursed in the UK under a risk-sharing scheme between the Department of Health and manufacturers. The plan included the cost of each of the disease-correcting treatments in the UK and included data collected from 2,048 MS patients, which was cost-effective across the country. the study identified the MS threshold as 36 36,000 per Quality-Adjusted Life Years (QALY). (12) Glatiramer acetate and interferon beta-1a 44 micrograms are currently the most widely prescribed drugs of any disease-modifying treatment on the US market. In 2013, the cost of medications for MS was estimated at $46.00 per member every year. This category of drugs is one of the second most expensive drugs for certain diseases. (13) In France, most remedial treatments in the field of disease correction for the treatment of relapsing-remitting multiple sclerosis are reimbursed. (14) Recently, oral therapies in France include teriflunomide in 2014 as a first-line drug, fingolimod in 2011 as a second-line drug, and delayed-release dimethyl fumarate in 2015 as a first and second-line drug introduced. With a new mechanism of action, dimethyl fumarate is an innovative approach in the treatment of relapsing-remitting multiple sclerosis. This oral medication provides patients with a comfortable treatment with the desired risk-benefit index. Dimethyl fumarate showed in phase three clinical trials to reduce the recurrence of multiple sclerosis by 53-44% and reduce the risk of three months of stable disability by 21-38%. (15-17) Also, Indirect comparisons showed that dimethyl fumarate is more effective than first-line teriflunomide injections, is as effective as fingolimod and is less effective than natalizumab in terms of disease recurrence rate. (18) Multiple sclerosis is especially characterized by the progression of the disease worse stages, with a high economic burden and reduced quality of life of patients. (17)

Multiple sclerosis affects the quality of life and performance of health-related patients, and the symptoms of this disease can place a significant economic burden on the patient, their family, health care system, and society at higher direct and indirect costs by increasing the severity of the disease. (19) Many attempts have been made to estimate the cost-effectiveness of these treatments for multiple sclerosis. (2) For this purpose, the present study aimed to analyze the cost-benefit and cost-effectiveness of relapsing-remitting drugs for multiple sclerosis.

## Materials&Methods

This review has analyzed the cost-utility and cost-effectiveness of disease-modifying drugs of Relapsing-Remitting Multiple Sclerosis via a systematic review-based approach. The review was conducted according to the Preferred Reporting Items for Systematic Reviews and Meta-Analyses (PRISMA) statement recommendations.

### Search strategy

We searched the electronic literature databases (Pubmed, Web of Science, Scopus, and Embase) with related keywords Including Disease-Modifying Drugs, Relapsing-Remitting Multiple Sclerosis, Cost-Utility Analysis, Cost-Effectiveness Analysis, Systematic Review (see supplementary material). The search strategy obeys the PICOS includes the following:

P: Includes all patients with multiple sclerosis and taking the drugs of relapsing-remitting Multiple Sclerosis (RRMS)

I: The disease-modifying drug of Relapsing-Remitting Multiple Sclerosis

C: Other types of drug and treatment methods that can be substituted.

O: Outcomes are reported based on ICER and costs per natural unit of health measurement.

S: Includes economic evaluation in a purely cost-effective manner.

### Inclusion Criteria

All studies that were published until 2019 examined the cost-utility and cost-effectiveness of disease-modifying drugs for patients with RRMS.

### Exclusion Criteria

In this study, articles were excluded that had the following criteria:

1. Items such as review articles, abstracts, protocols, letters to the editor, etc.
2. Articles whose full text has been published other than in English.
3. Studies for which full text was not available. Google Scholar sites were also used to complete the search.

Studies that performed incomplete economic evaluation (effectiveness evaluation, cost evaluation), studies that were in the form of reviews, conference abstracts, letters to the editor and protocol, and studies that were not in English and Persian languages, had to be excluded.

### Data collection and analysis

#### Selection of studies

All studies related to Cost-Utility and Cost-Effectiveness Analysis of RRMS were selected according to the inclusion and exclusion criteria mentioned above and duplicate cases were removed using Endnote 20 software. In the first stage, the title and summary of the remaining studies were studied independently by two researchers. If there was a disagreement, the third researcher reviewed the studies to avoid any bias. In the next stage, the full text of the studies was carefully examined by two researchers separately. Any disagreements between the researchers were addressed by the third person.

#### Synthesis

Accurate information from each study was obtained separately using a data extraction form pre-made by the two authors. Background information about the study included the author’s name, year of publication, outcome measure, setting, study population, interventions, type of economic evaluation, perspective, time horizon, willingness to pay (WTP) threshold, discount rate, sensitivity analyses, etc. Moreover, To compare costs in different studies, all costs used were converted to 2020 dollars based on Purchasing Power Parity (PPP).

#### Quality assessment

The PRISMA checklist was used to assess the quality of this study. More details about quality assessment are reported in Table 1.

**Table 1.**
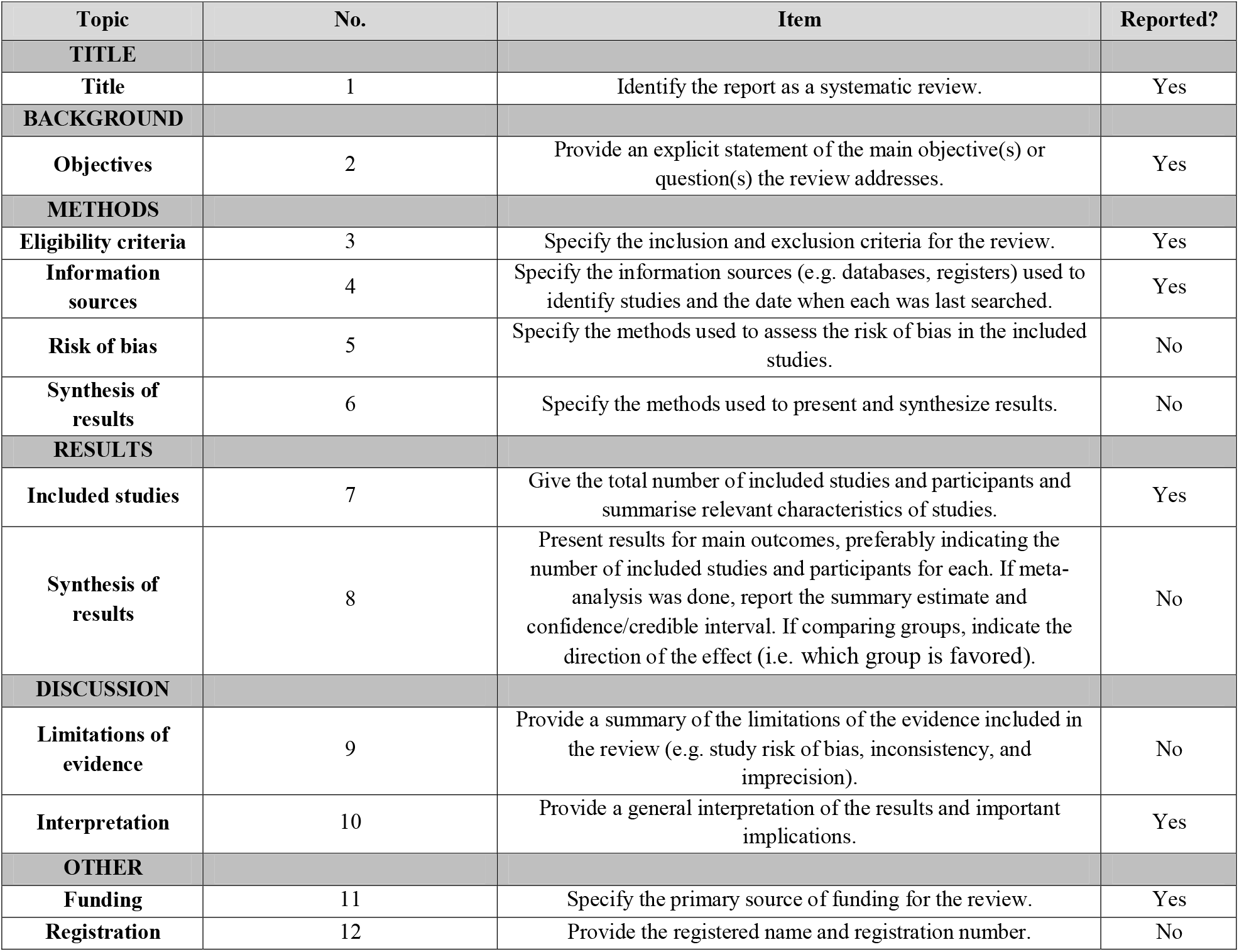
Evaluate this article based on the PRISMA checklis

## Results

The initial search results produced 1,360 records, 469 studies were omitted due to duplication between databases and other information sources. According to the inclusion and exclusion criteria of the study, 513 studies in the title study, 263 studies in the abstract review, and 94 studies in the full-text review were omitted. Based on the PRISMA flowchart in Figure 1, which shows the process of reviewing and selecting studies, finally, 21 studies were included in the review.

**Figure 1.**
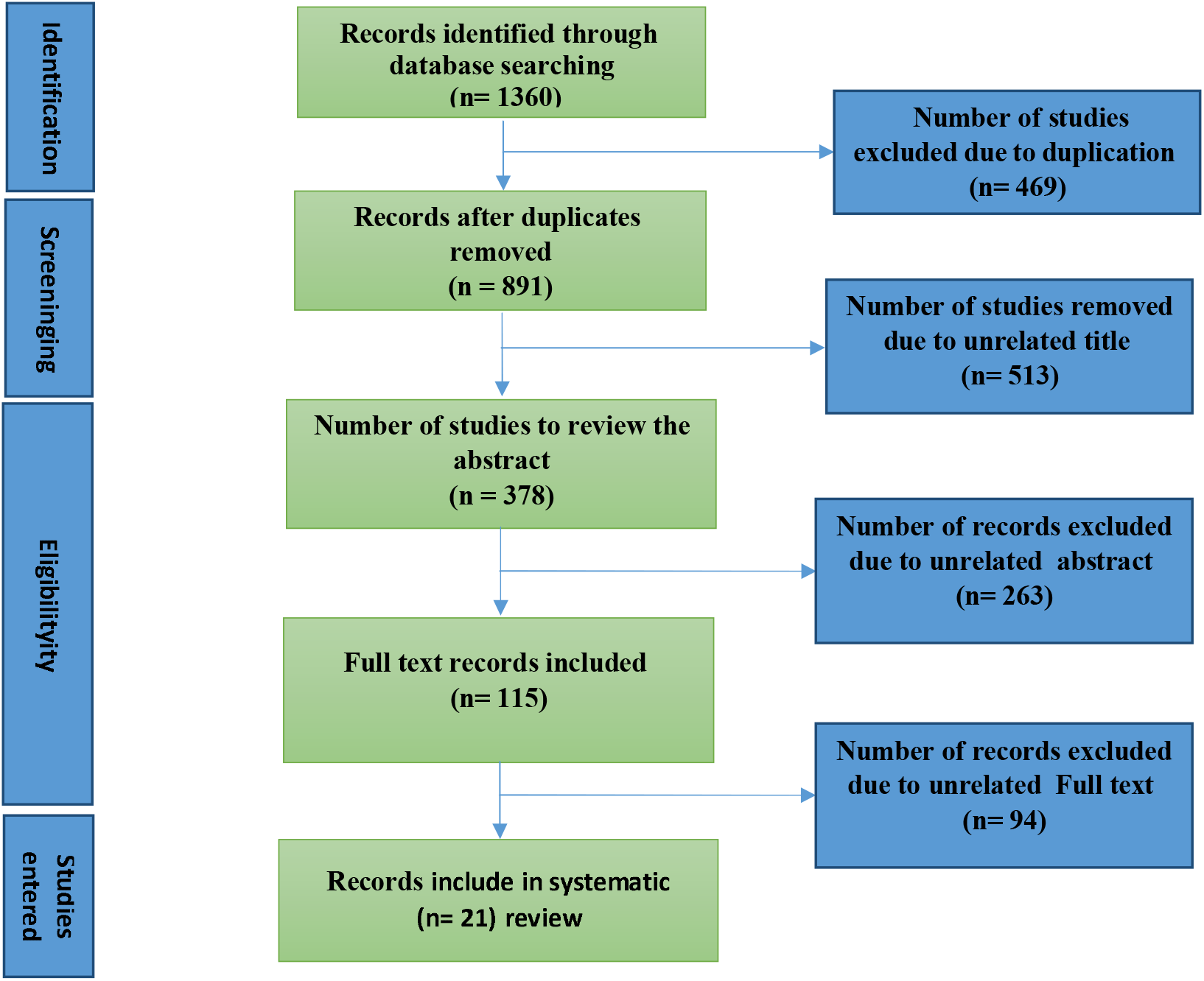
PRISMA flowchart

### Study characteristics (see Table 2)

#### Location

Ten articles were published in European countries, four in the United States, two in Canada, three in Iran, and finally one in Thailand and Saudi Arabia.

**Table 2.**
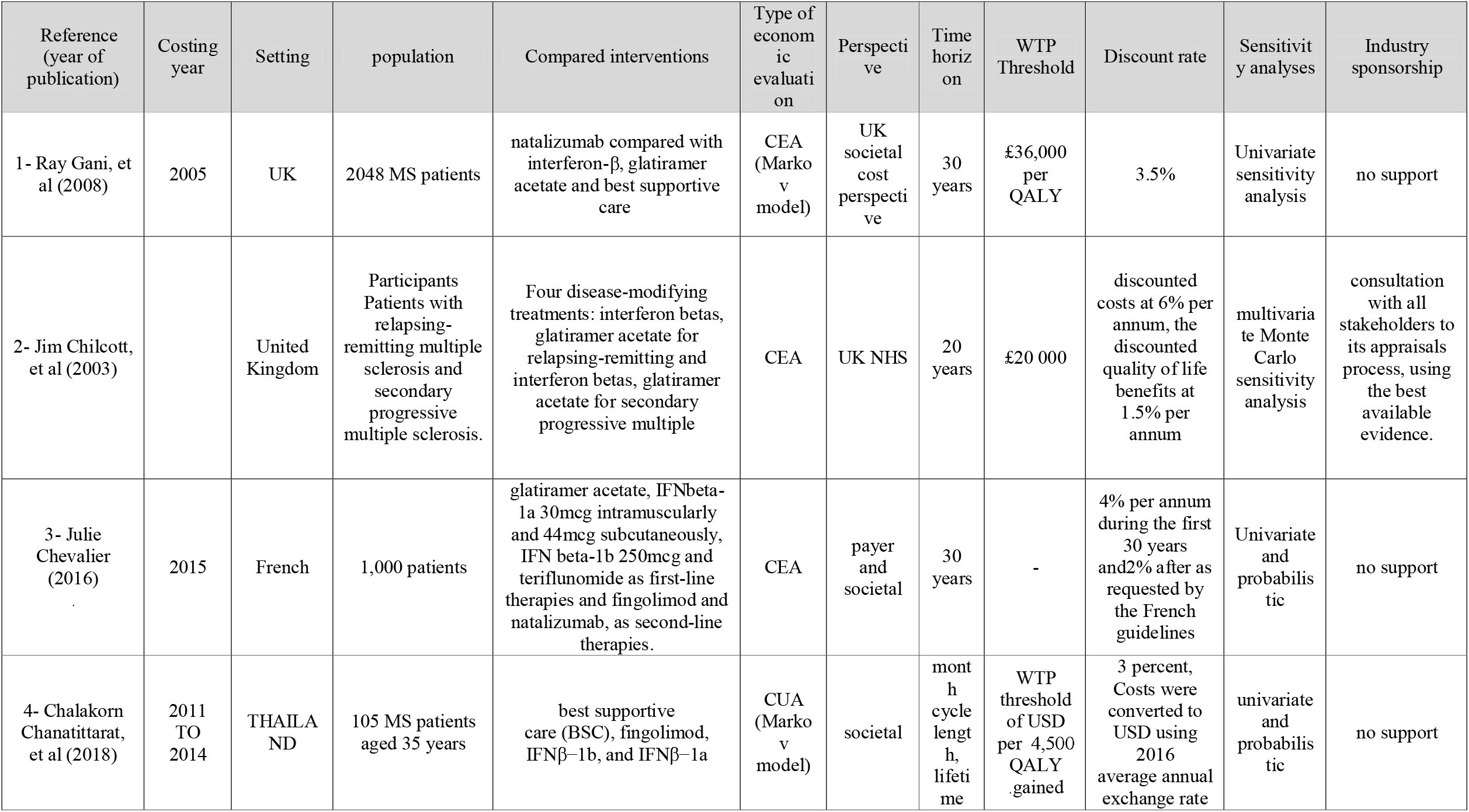
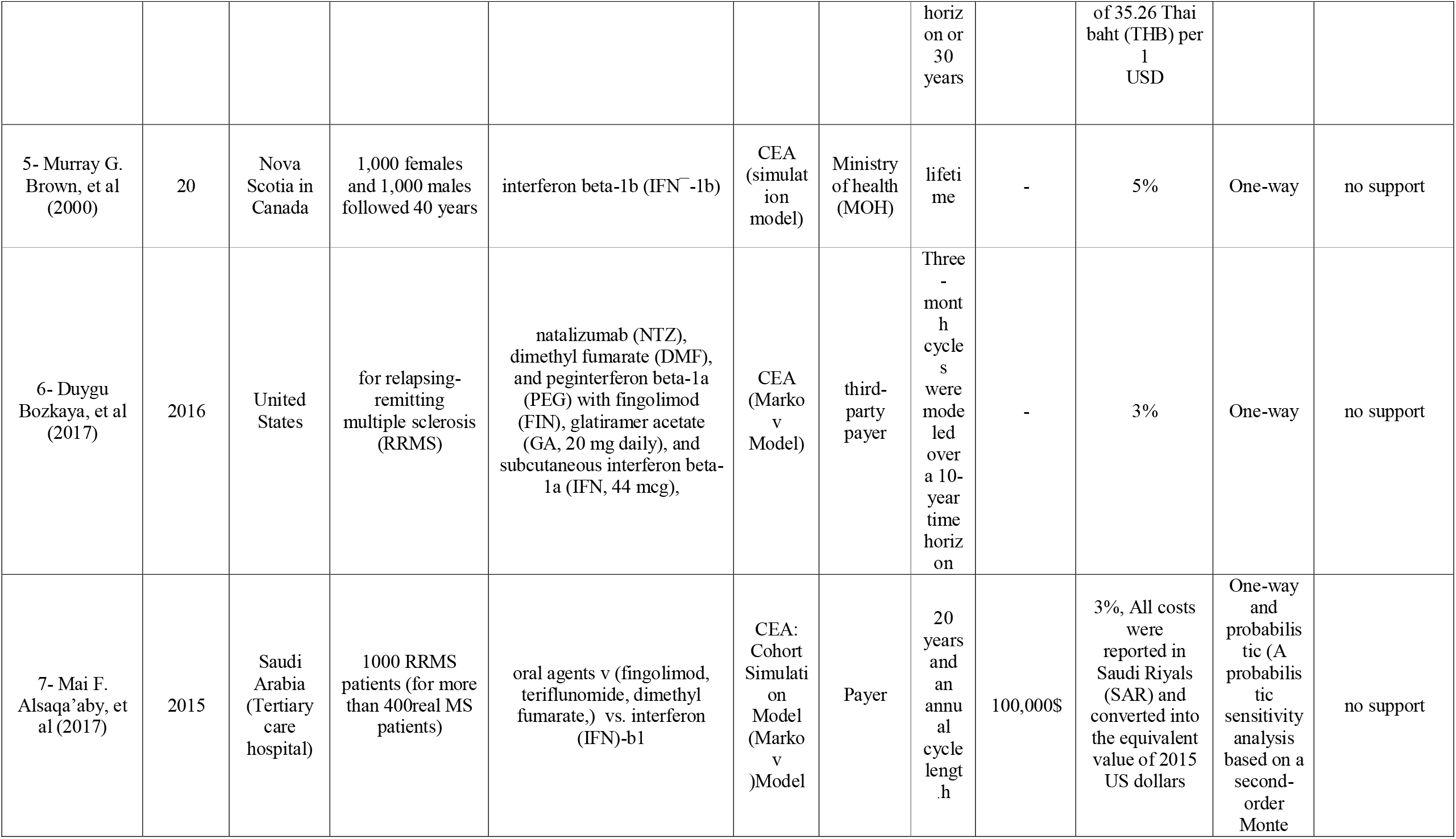
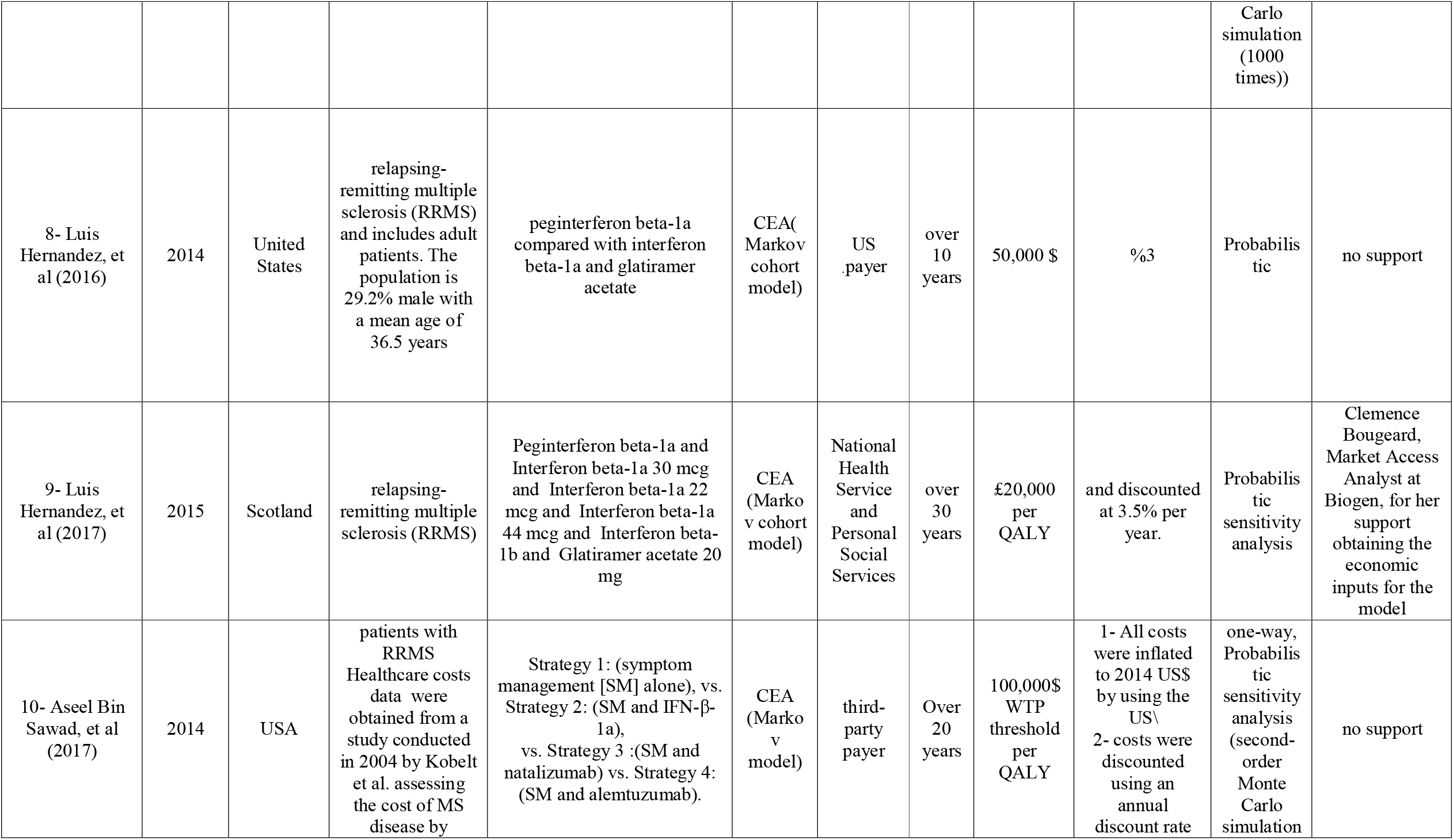
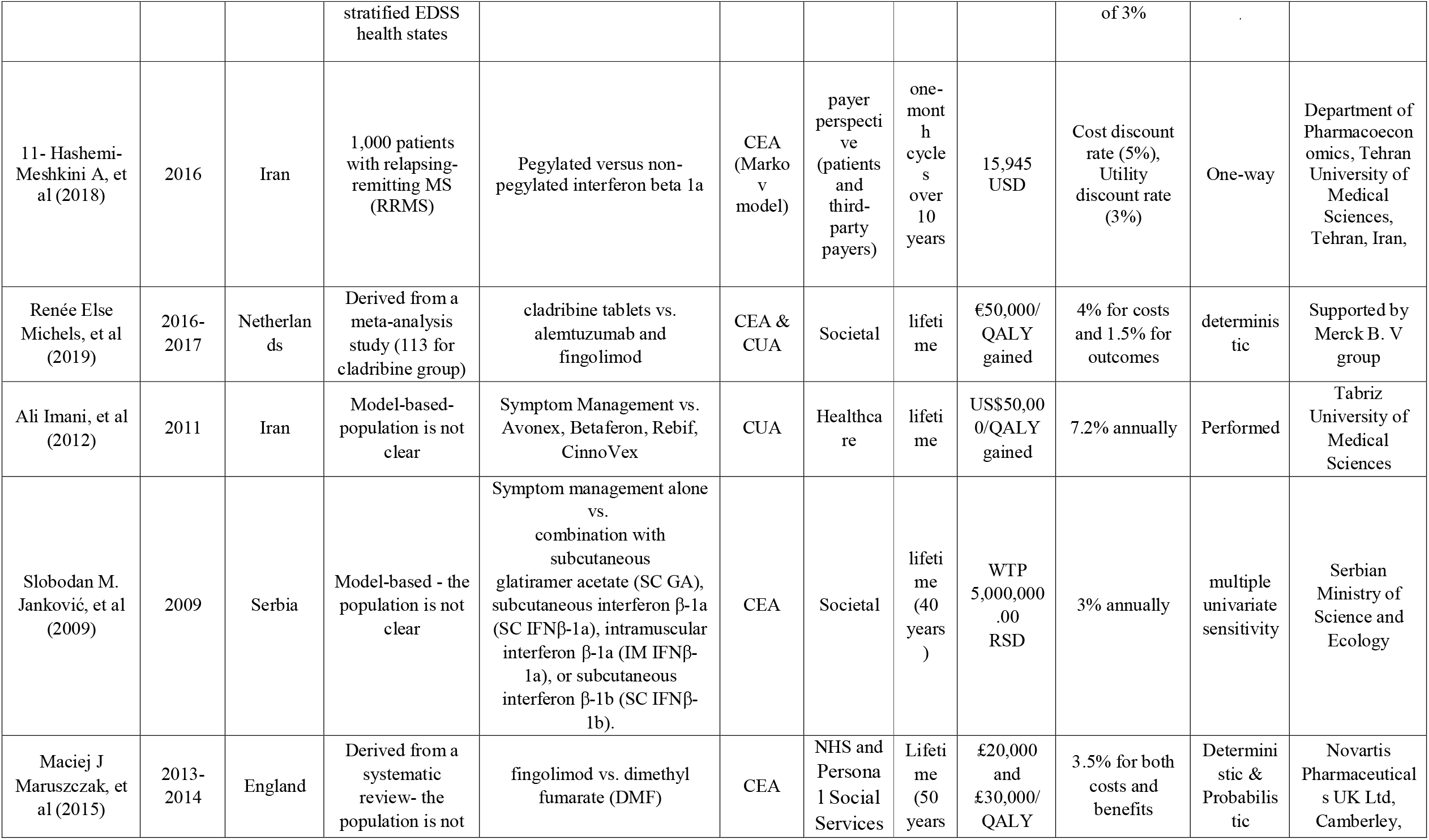
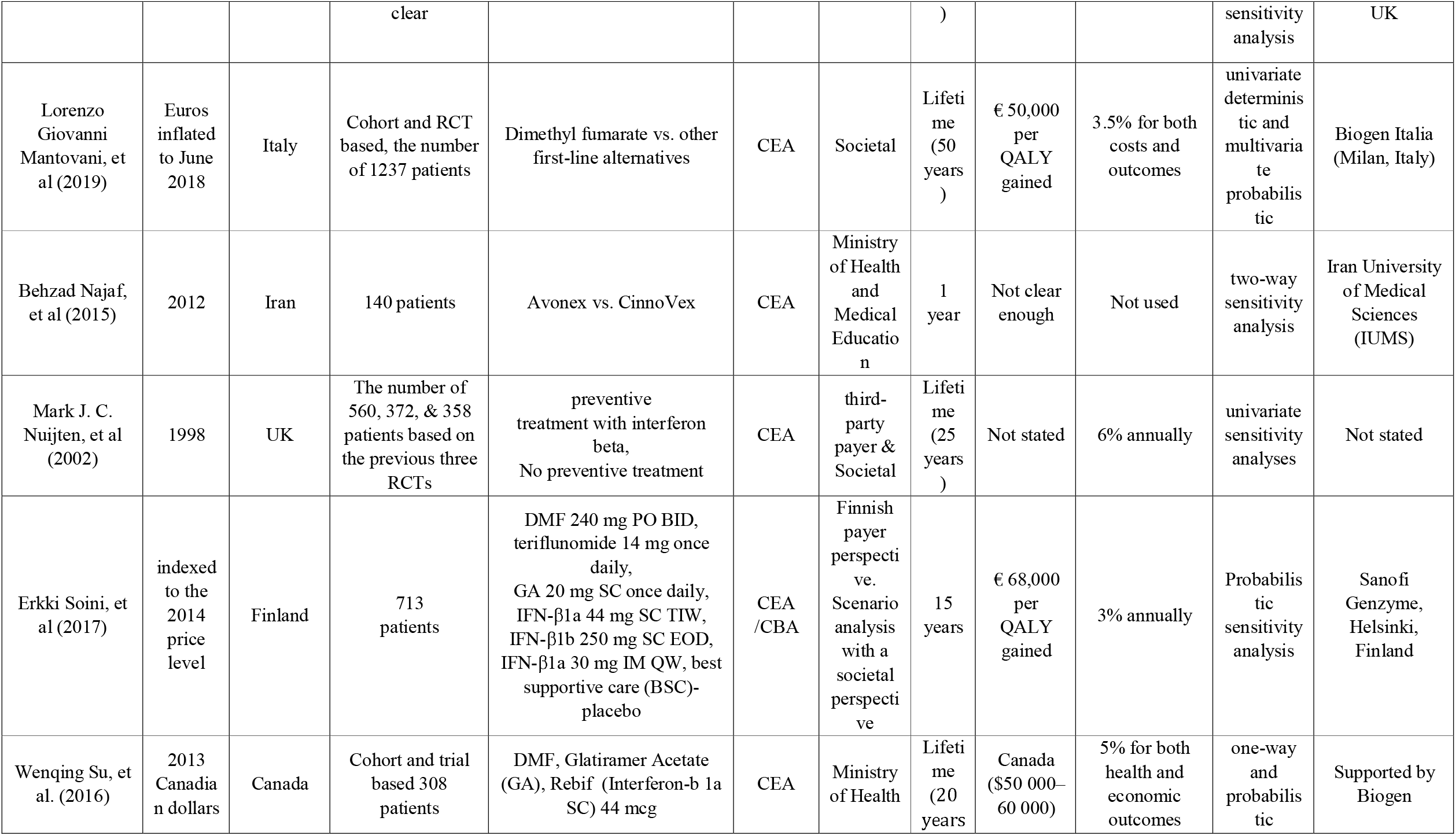
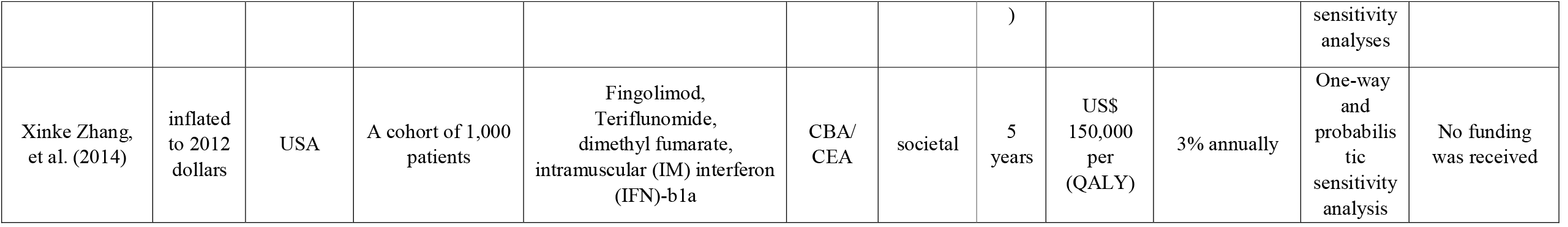
Study characteristics

#### Type of economic evaluation

The review of the final studies showed that seventeen articles specifically analyzed the cost-effectiveness of disease-modifying drugs. Two articles analyzed the cost-effectiveness of these drugs and two articles combined the cost-benefit and cost-effectiveness of disease-modifying drugs.

#### Perspective

there were three articles from the patient’s perspective, eight articles from the health system’s perspective, and seven articles from the community’s perspective. In addition, the perspective of an article was a combination of patient and health system, the perspective of an article was a combination of community and health system and the perspective of an article was a combination of patient and community.

#### Sensitivity analysis method

To explain in terms of sensitivity analysis, seven articles used one-way methods, and one article used two-way sensitivity analysis. Also, two articles used deterministic sensitivity analysis (DSA) and eleven articles used probabilistic sensitivity analysis (PSA). Also, in terms of several variables, three articles used a multiple univariate sensitivity method and five articles used the univariate sensitivity method.

#### Forms of disease-modifying drugs of RRMS

Three forms of treatment of RRMS were identified; injectable, oral, and intravenous infusions treatments. Eight studies only analyzed the injectable disease-modifying drugs of RRMS together (2, 8, 13, 20-24). Two studies only analyzed the oral disease-modifying drugs of RRMS together (25, 26). Four studies analyzed the injectable and oral disease-modifying drugs of RRMS (27-30). Of course, in one study, in addition to these two forms of medication, the best supportive care (BSC) strategy was included in cost-effectiveness analysis (31).

Three studies analyzed the injectable and intravenous infusions disease-modifying drugs of RRMS. Of course, in all three studies, symptom management was one of the strategies analyzed along with the two strategies of injectable drugs and intravenous infusion (12, 32, 33).

One study analyzed the oral and intravenous infusions disease-modifying drugs of RRMS (34). Finally, in two studies, all three types of disease-modifying drugs of RRMS have been analyzed together (15, 35).

#### Outcome and Cost (see Table 3)

The results reported in the studies mainly include incremental cost and incremental cost-effectiveness ratio (ICER) per different natural units particularly the quality of life years (QALYs). The lowest and highest numerical value of outcome measures were -1,623,918 and 2,297,141.53, resprectively. In article of Sawad et al., the lowest numerical value was related to the comparison of strategies 4 (symptom management (SM) and alemtuzumab) and 3 (SM and natalizumab). Also, In the same article, the highest numerical value was related to the comparison of strategies 2 (symptom management (SM) and IFN-β-1a) and 1 (SM alone) (32). The lowest numerical value of the cost of disease-modifying drugs of RRMS was $180.67 for total cost of Natalizumab in first year considered in the article (34), the highest value of cost was $1474840.19 for the total lifetime cost per patient treated with IFN beta-1b -250 mcg (26).

**Table 3.**
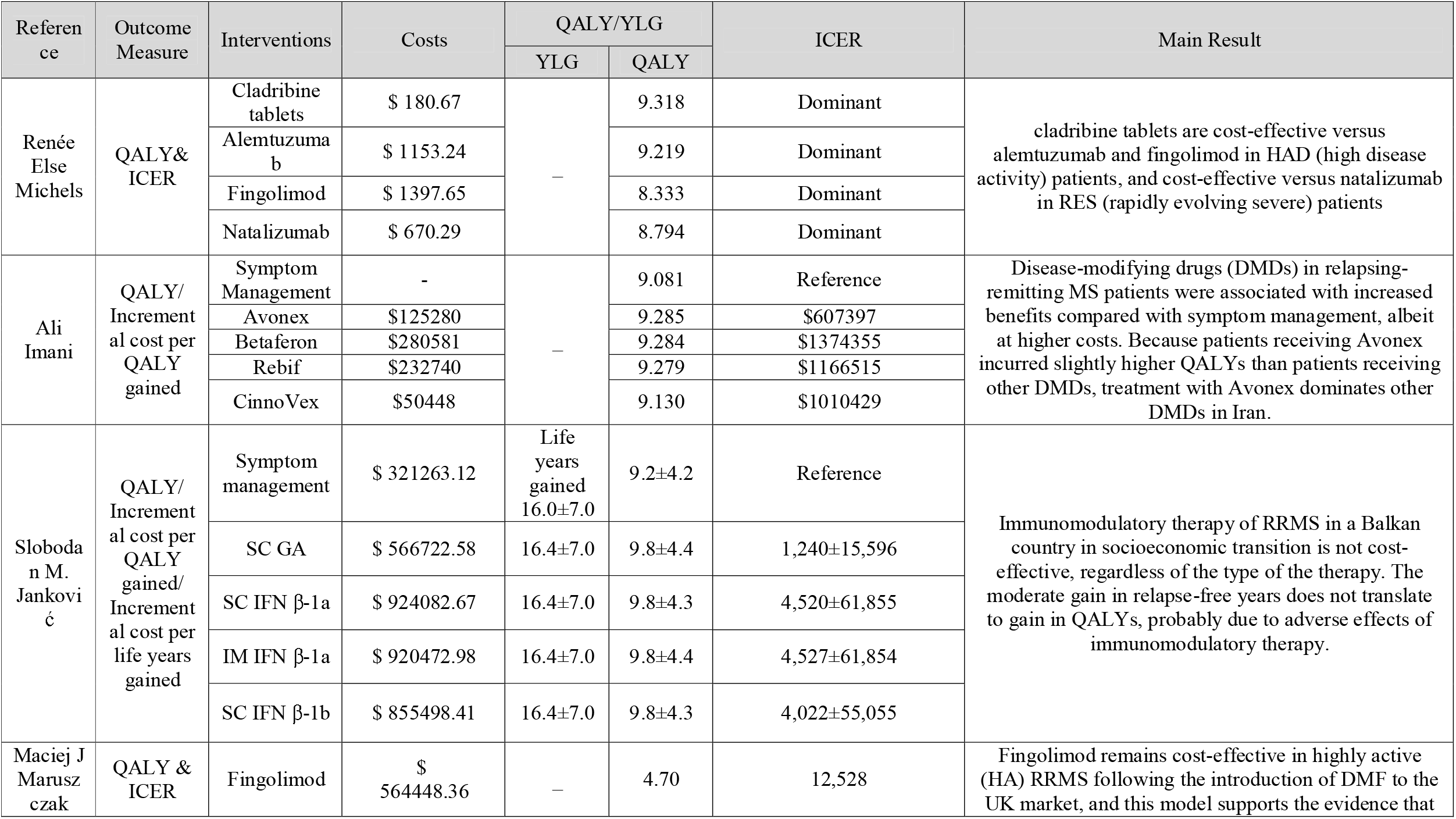
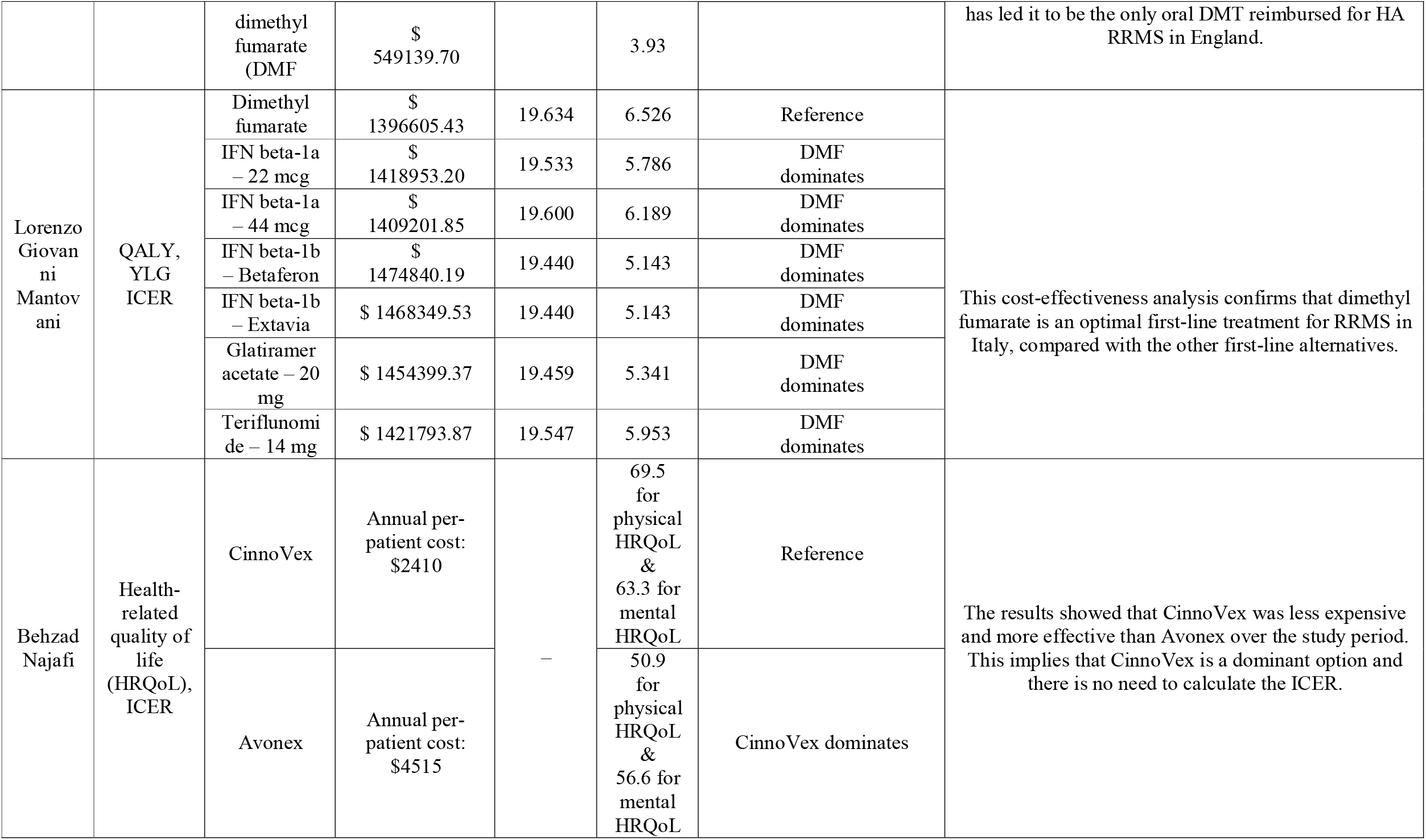
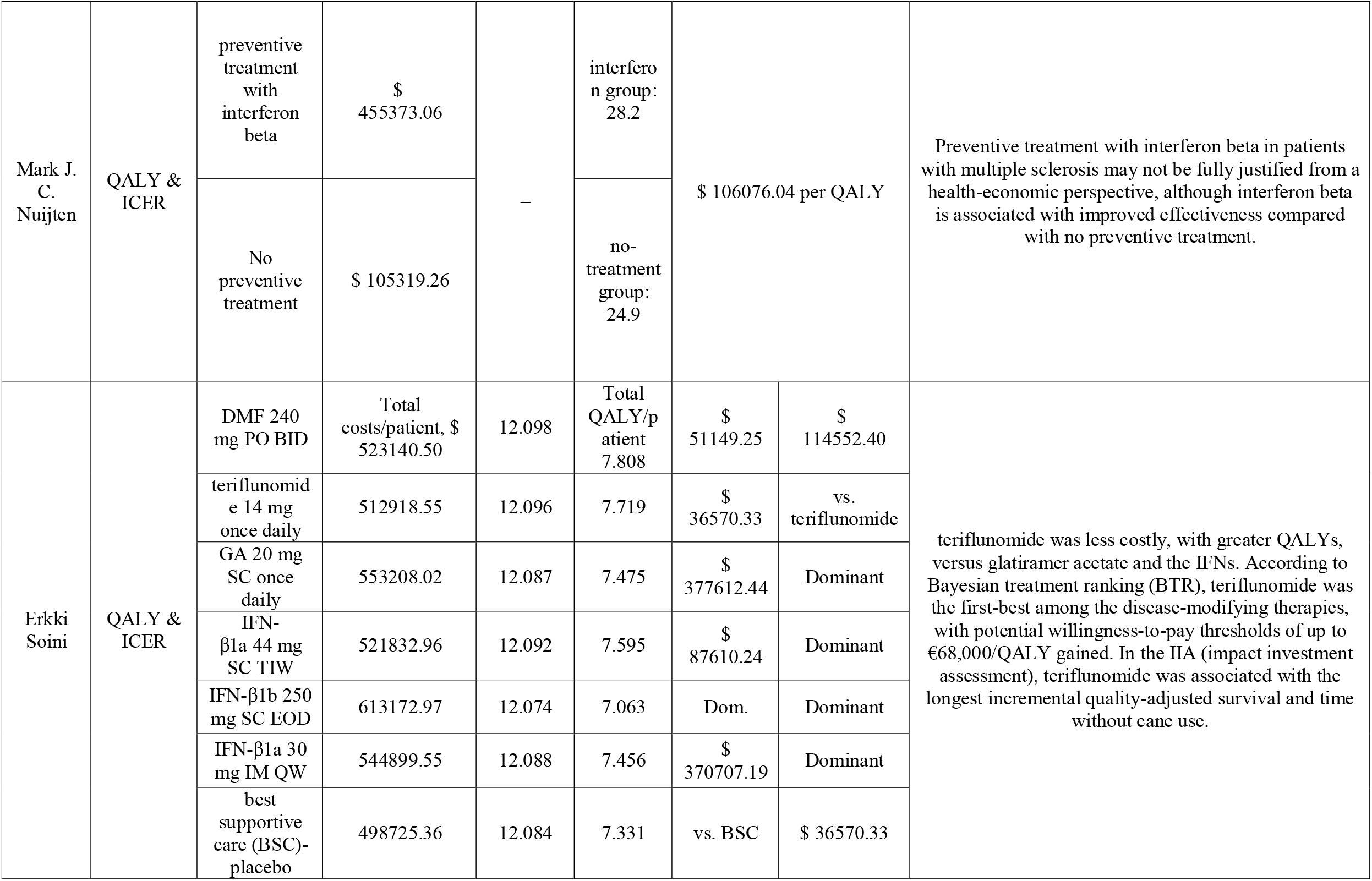
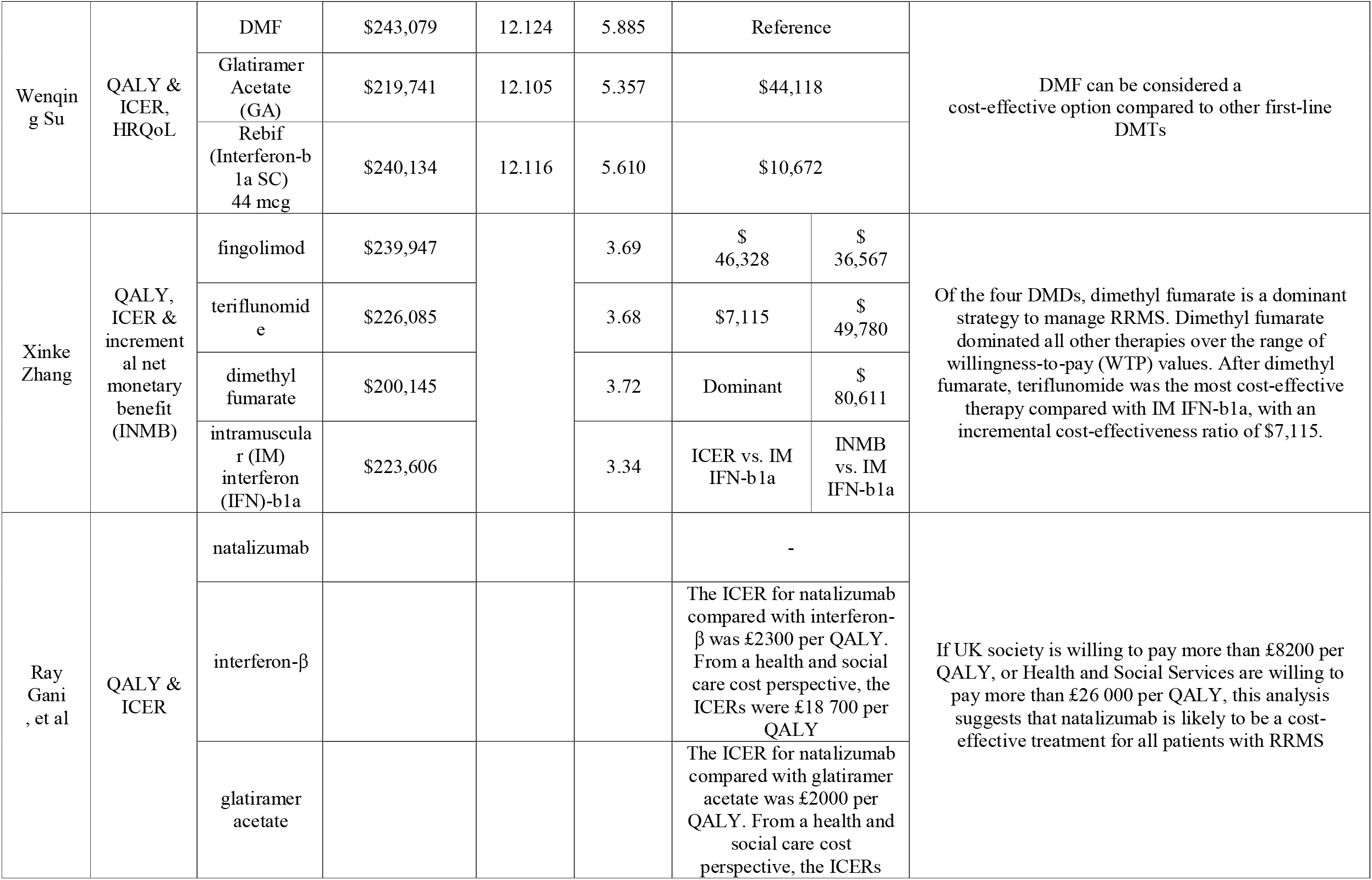
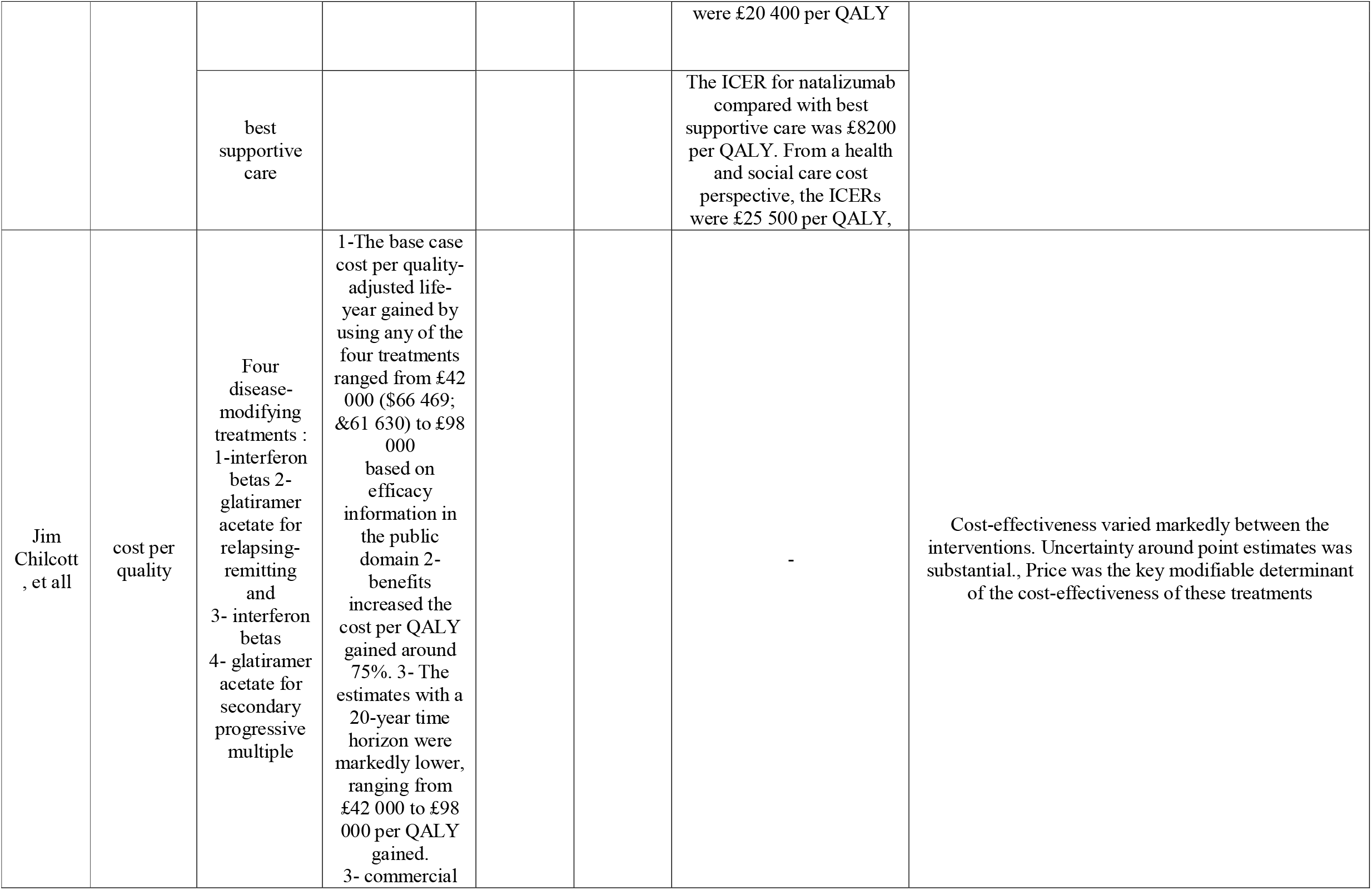
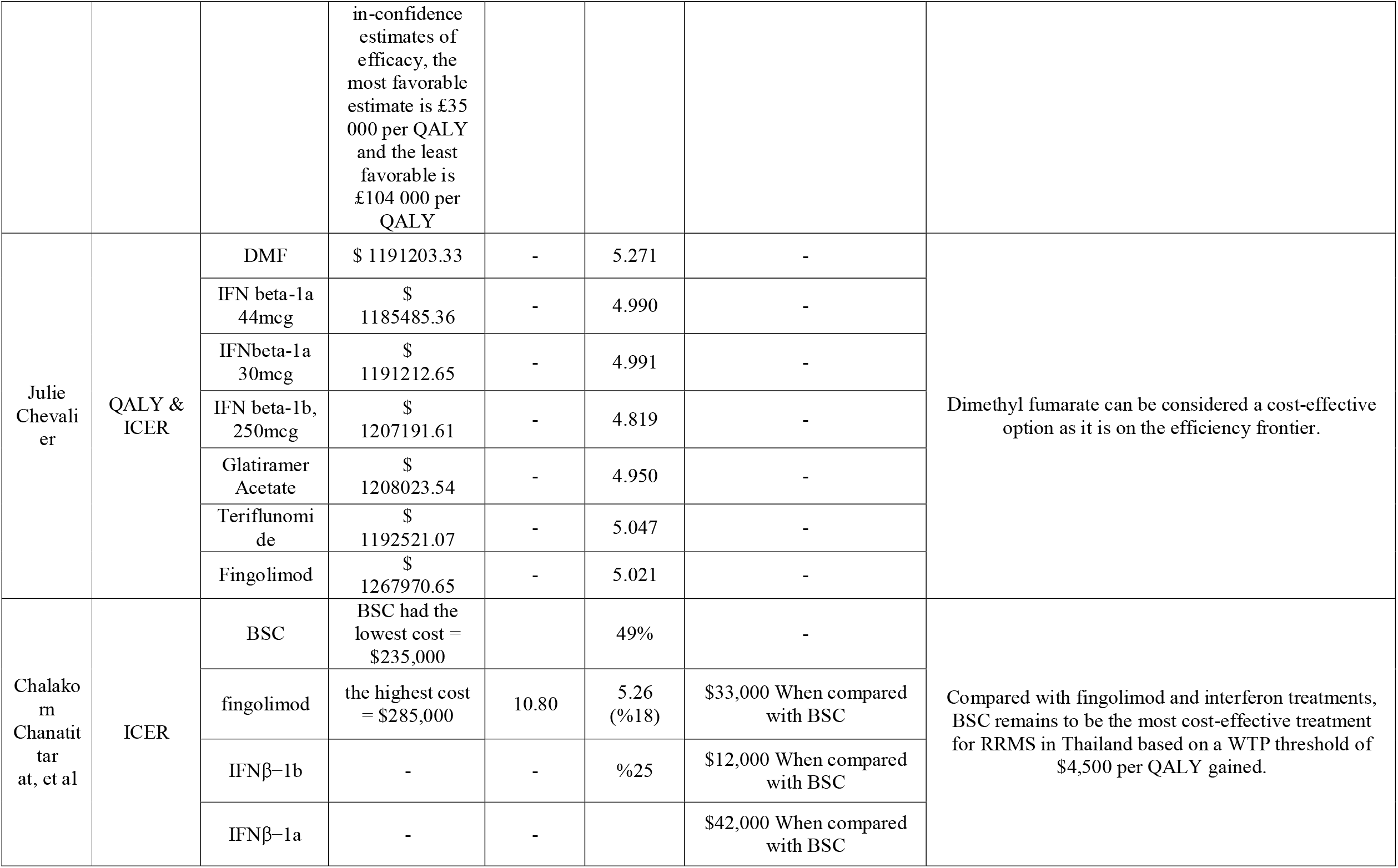
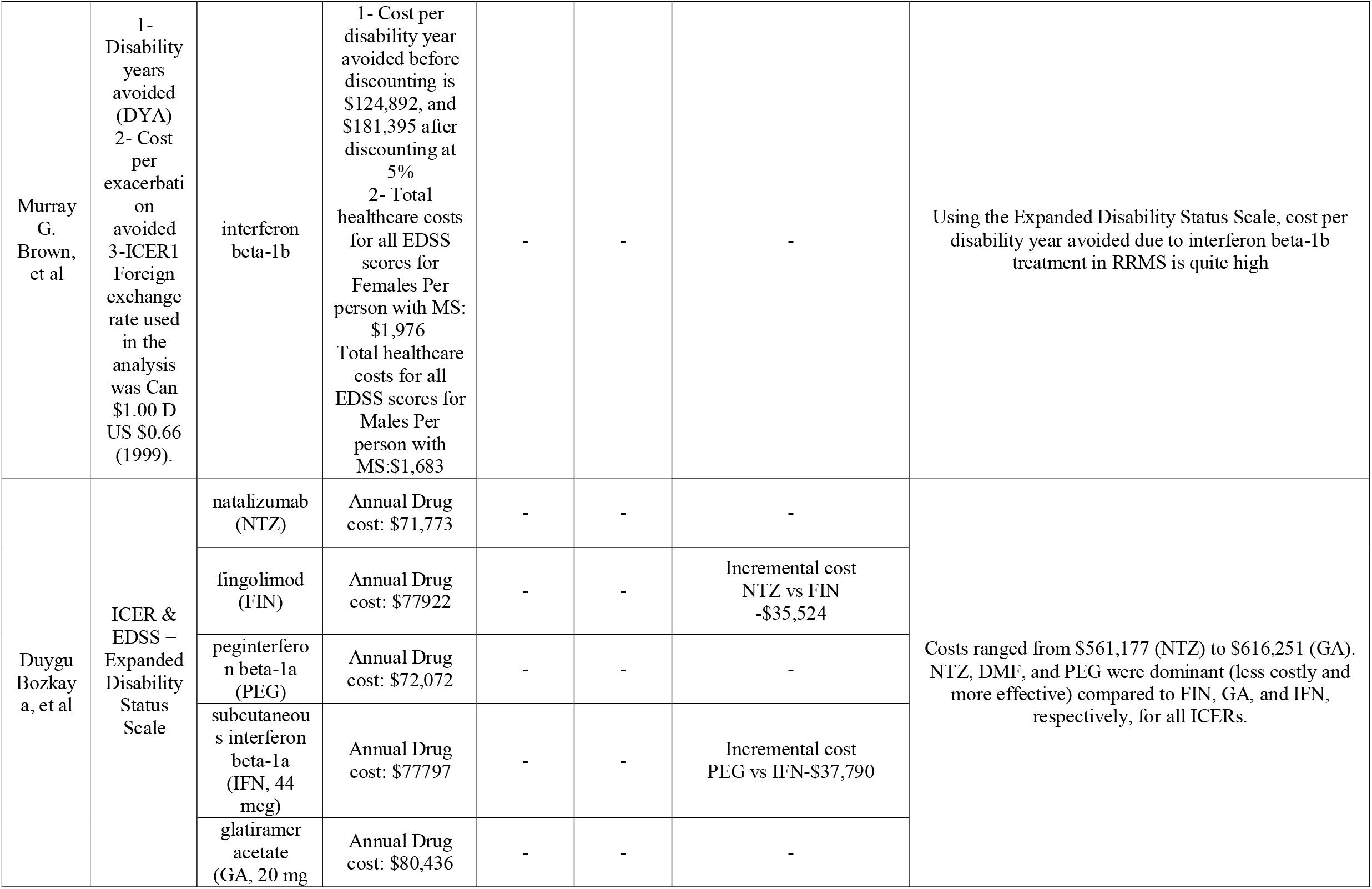
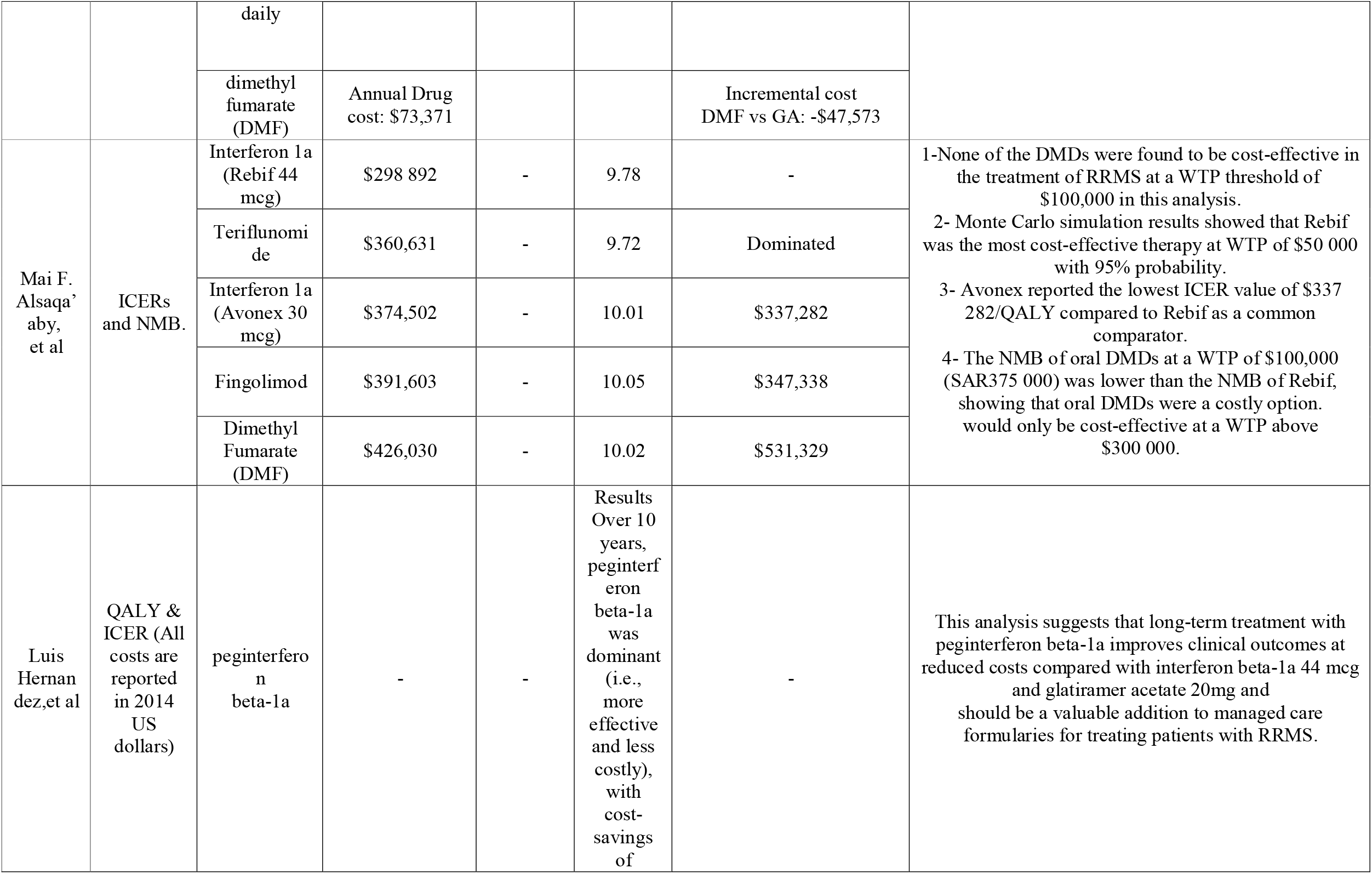
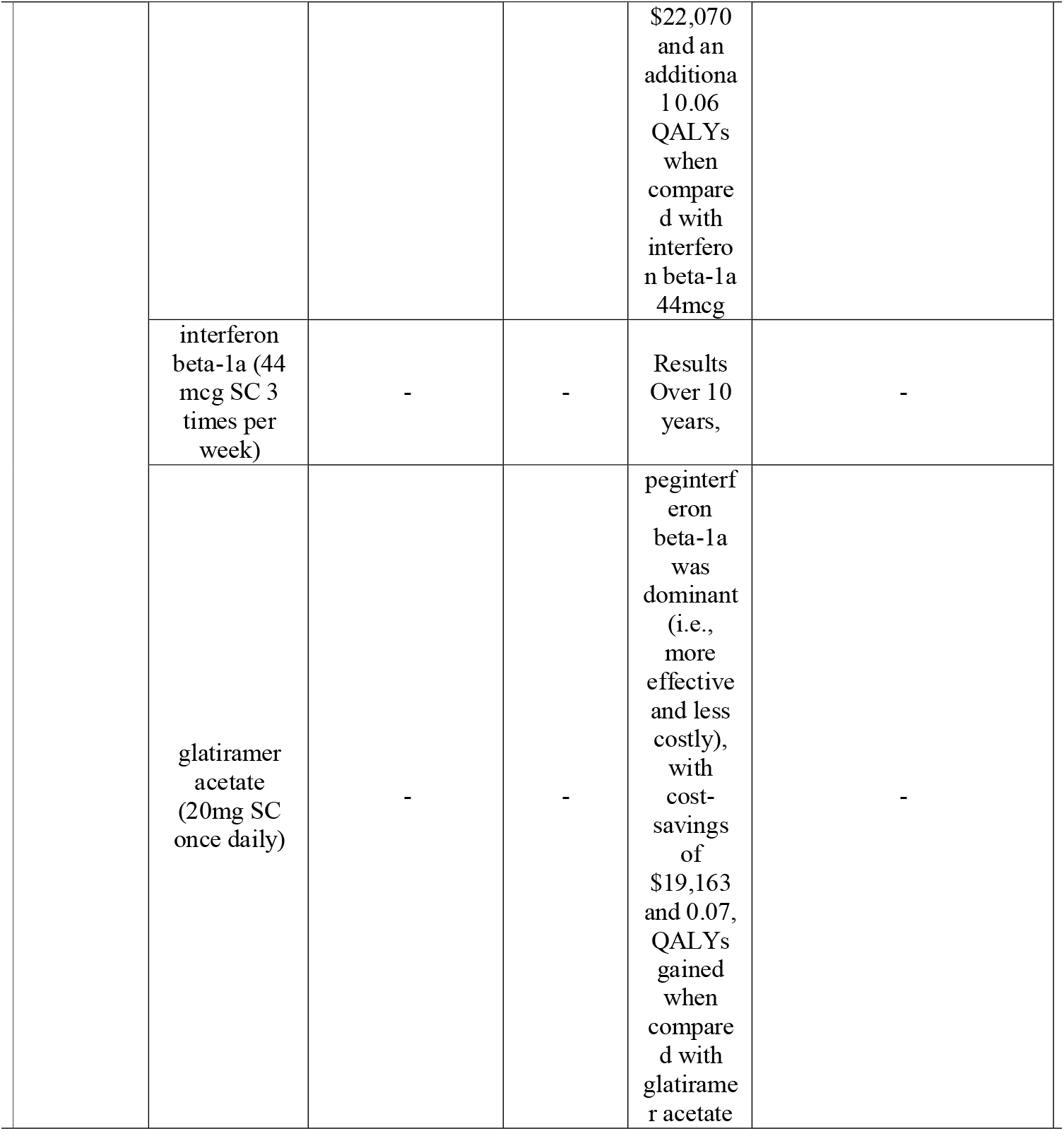
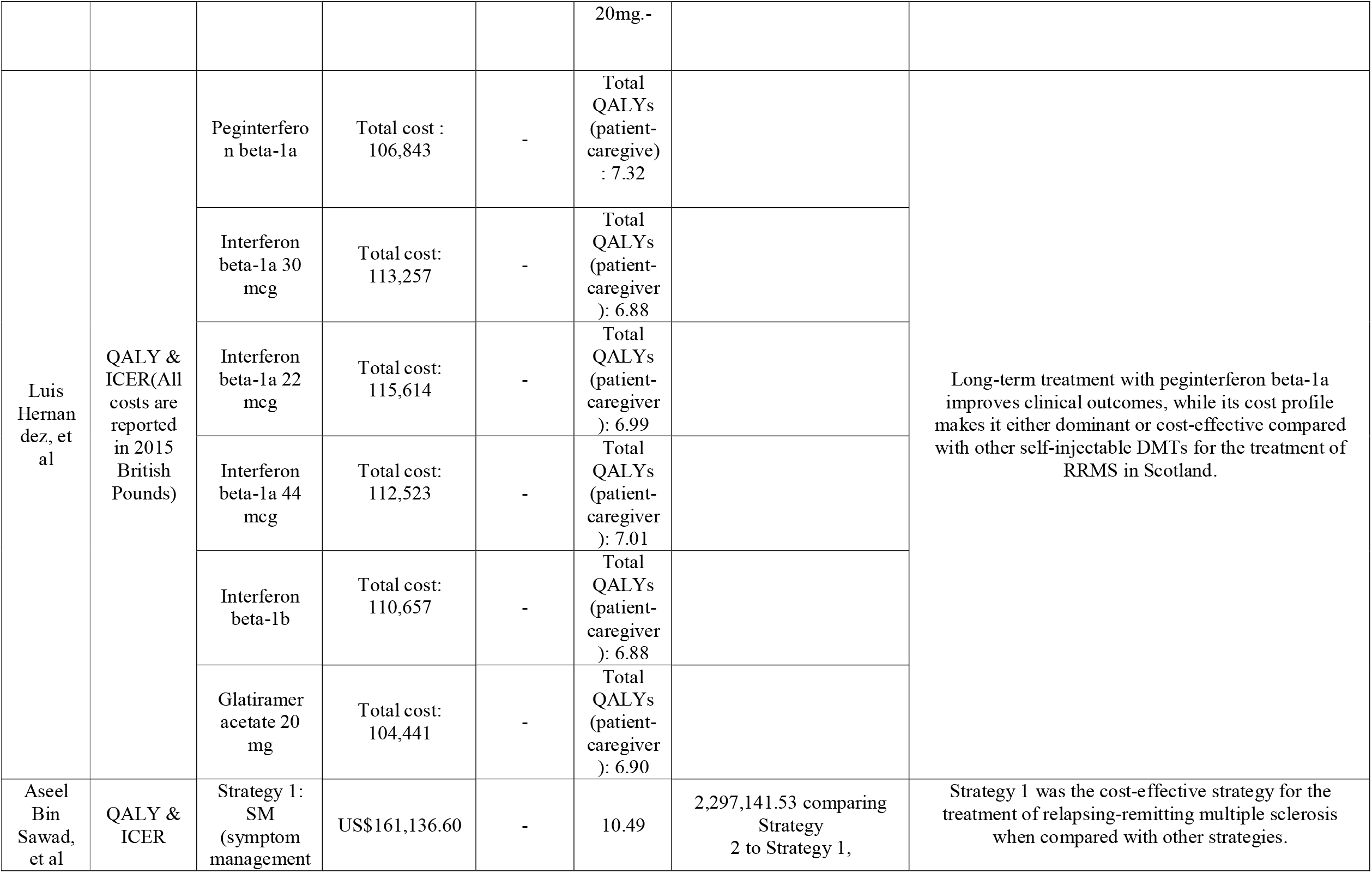
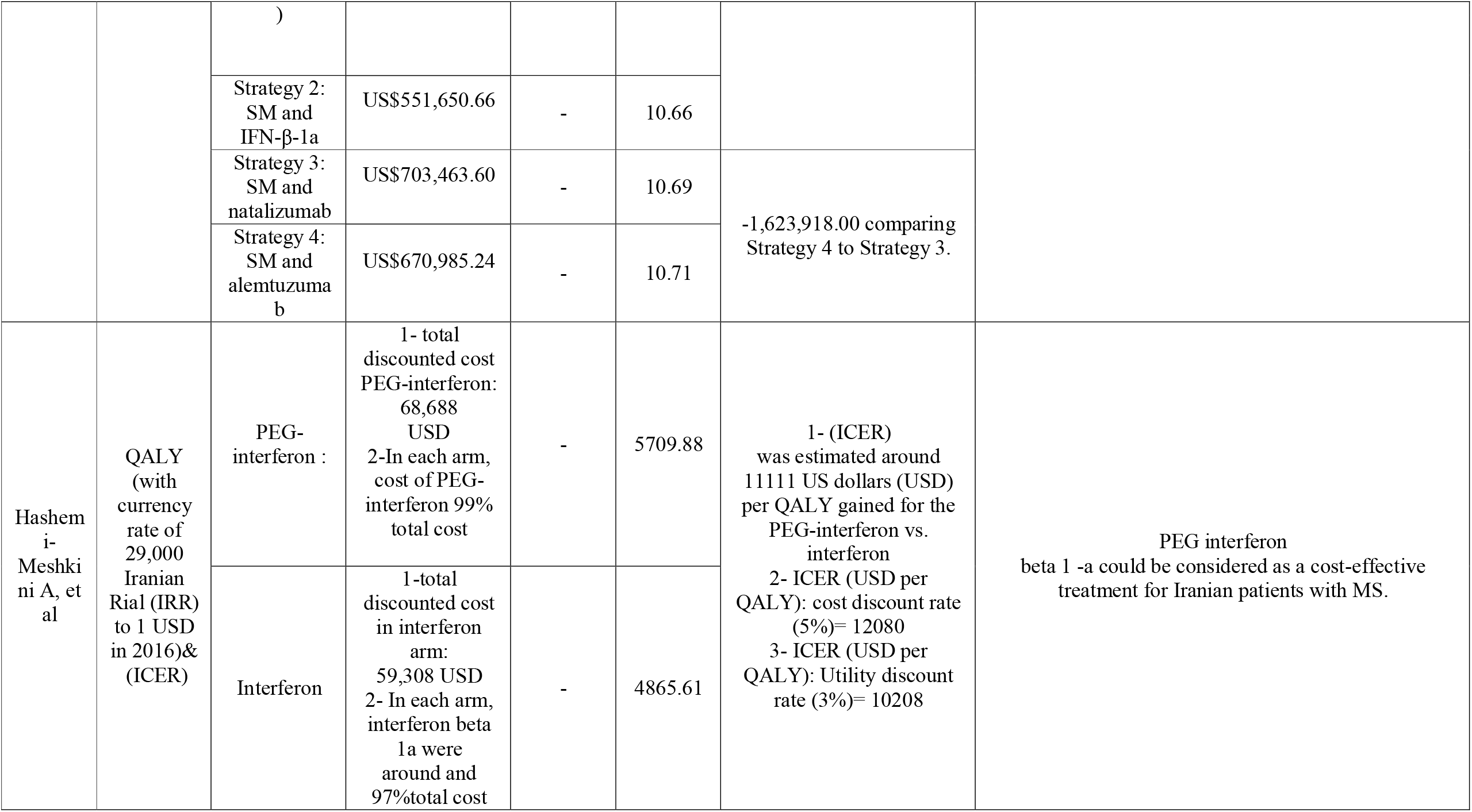
Outcome and Cost

## Discussion

This is the first systematic review to examine the Cost-Utility and Cost-Effectiveness Analysis of disease-modifying drugs of RRMS. In this article, twenty-one studies were included in the review. The disease-modifying drugs of RRMS can be divided into injectable, oral, and intravenous infusions. The most important injectable disease-modifying drugs of RRMS are interferon beta-1a (Avonex and Rebif) and beta-1b (Betaferon and Extavia), glatiramer acetate (Copaxone), ofatumumab, etc. Also, the most important oral disease-modifying drugs of RRMS are teriflunomide (Aubagio), monomethyl fumarate (Tecfidera), fingolimod, cladribine, siponimod, ponesimod, dimethyl fumarate (DMF), diroximel fumarate, ozanimod, etc. Finally, the most important intravenous infusions disease-modifying drugs of RRMS are alemtuzumab, mitoxantrone, ocrelizumab, natalizumab, etc.

Some disease-modifying drugs such as Avonex, Rebif, Plegridy, Betaferon, Copaxone, Extavia, Tecfidera, and Aubagio are considered as first-line treatments of patients diagnosed with RRMS (15, 36, 37). In some articles, such as Imani et. al., in addition to symptom management strategy, the cost-effectiveness analysis of four first-line drugs, including Avonex, Betaferon, Rebif, and CinnoVex are also analyzed (22). Two disease-modifying drugs Included fingolimod and natalizumab are considered as second-line treatments of patients diagnosed with RRMS (36, 37). Also, in an article such as Bozkaya, et al in addition to drugs such as peginterferon beta-1a, IFN, 44 mcg, glatiramer acetate, 20 mg, and DMF, the cost-effectiveness analysis of two second-line drugs, including natalizumab and fingolimod are analyzed (35). Finally, some drugs such as alemtuzumab (Lemtrada) and mitoxantrone (Ralenova) are formally second-line drugs but are prescribed as third-line drugs (37).

In general, in evaluating the cost-effectiveness of the three forms of disease-modifying drugs of RRMS, the oral form of these drugs was more effective than the two forms of injection and intravenous infusion. For example, Julie Chevalier and colleagues analyzed drugs in injectable, oral, and intravenous infusions together and concluded that the oral drug DMF was more cost-effective than injectable and intravenous infusions. In this study, ICER DMF was $ 20,348.15 per QALY compared to IFN beta-1a 44mcg injection; Of course, from the payer’s point of view, this amount was equivalent to $ 45084.11 per QALY. The univariate analysis confirmed that this drug was the most cost-effective compared to other drugs studied (36).

Four studies analyzed the cost-effectiveness of injectable disease-modifying drugs of RRMS with the oral form of these drugs. According to these studies, in three studies, oral drugs were more effective than injectable drugs. For example, a study by Xinke Zhang et al. analyzed the cost-effectiveness of fingolimod, teriflunomide, dimethyl fumarate, and intramuscular (IM) interferon (IFN) -b1a, found that the cost-effectiveness of dimethyl fumarate compared to other There are more drugs. Other than dimethyl fumarate, teriflunomide is cheaper than IM IFN-b1a with ICER (the US $ 7,115). However, the results in this study are reported in terms of incremental net monetary benefit (INMB). In a study by Wenqing Su et al., Which analyzed the cost-effectiveness of Dimethylformamide (DMF), Glatiramer Acetate (GA), and Rebif Interferon-b 1a), it was reported that DMF was more cost-effective than other drugs compared.. In general, oral medications are preferred by patients due to their non-invasive nature compared to other forms of medication. Also, they generally have lower costs, which plays a significant role in their greater cost-effectiveness. Also, in a study, the cost-effectiveness of these two drug forms with BSC has been analyzed and the cost-effectiveness of the BSC strategy has been concluded; One reason for the cost-effectiveness of this strategy is the lower cost of medicine and equipment, as well as the greater effectiveness of the best patient care. In two studies, the cost-effectiveness of injectable drugs and intravenous infusions was analyzed by symptom management; In both studies, the cost-effectiveness of symptom management was higher compared to the other two strategies, which can be attributed to the lower cost of drugs and equipment (32, 33). Some published studies, such as Prosser et al. And Rubio-Terres et al., Have analyzed the cost-effectiveness of drug forms with BSC (38, 39).

Three studies analyzed the cost-effectiveness of injecting drug-modifying drugs of RRMS, intravenous infusions, and symptom management of RRMS. In two studies, symptom management was more effective, and in one study, drugs in the form of intravenous infusions were more effective. In this study, to help calculations and increase the validity of the results obtained from various articles, some measures were taken such as converting different currencies into one currency.

Moreover, in this systematic review study based on the designed research approach, only studies published in English were considered.

## Conclusion

Based on the results of all studies, it can be concluded that for the treatment of patients with AF, care-oriented strategies such as BSC and SM strategies should be preferred to drug strategies due to their greater cost-effectiveness, and policies and planning should be to implement this strategy more widely and with better quality. Also, among the drug strategies with different prescribing methods, oral disease-modifying drugs of RRMS should be preferred to injectable drugs and intravenous infusions for various reasons such as their non-invasiveness and greater convenience for patients, and lower cost.

## Data Availability

All data produced in the present work are contained in the manuscript

## Acknowledgments

The authors appreciate the cooperation of the Health Management and Economics Research Center at the Iran University of Medical Sciences.

