## Supplementary material for "Cost-Utility and Cost-Effectiveness Analysis of disease-modifying drugs of Relapsing-Remitting Multiple Sclerosis: A Systematic Review": Table 3

| **Table 3. Outcome and Cost** | | | | | | | | | |
| --- | --- | --- | --- | --- | --- | --- | --- | --- | --- |
| Reference | Outcome Measure | Interventions | Costs | QALY/YLG | | ICER | | | Main Result |
|  |  |  |  | YLG | QALY |  |  |  |  |
| Renée Else Michels | QALY& ICER | Cladribine tablets | $ 180.67 | _ | 9.318 | Dominant | | | cladribine tablets are cost-effective versus alemtuzumab and fingolimod in HAD (high disease activity) patients, and cost-effective versus natalizumab in RES (rapidly evolving severe) patients |
|  |  | Alemtuzumab | $ 1153.24 |  | 9.219 | Dominant | | |  |
|  |  | Fingolimod | $ 1397.65 |  | 8.333 | Dominant | | |  |
|  |  | Natalizumab | $ 670.29 |  | 8.794 | Dominant | | |  |
| Ali Imani | QALY/ Incremental cost per QALY gained | Symptom Management | - | _ | 9.081 | Reference | | | Disease-modifying drugs (DMDs) in relapsing-remitting MS patients were associated with increased benefits compared with symptom management, albeit at higher costs. Because patients receiving Avonex incurred slightly higher QALYs than patients receiving other DMDs, treatment with Avonex dominates other DMDs in Iran. |
|  |  | Avonex | $125280 |  | 9.285 | $607397 | | |  |
|  |  | Betaferon | $280581 |  | 9.284 | $1374355 | | |  |
|  |  | Rebif | $232740 |  | 9.279 | $1166515 | | |  |
|  |  | CinnoVex | $50448 |  | 9.130 | $1010429 | | |  |
| Slobodan M. Janković | QALY/ Incremental cost per QALY gained/ Incremental cost per life years gained | Symptom  management | $ 321263.12 | Life years gained 16.0±7.0 | 9.2±4.2 | Reference | | | Immunomodulatory therapy of RRMS in a Balkan country in socioeconomic transition is not cost-effective, regardless of the type of the therapy. The moderate gain in relapse-free years does not translate to gain in QALYs, probably due to adverse effects of immunomodulatory therapy. |
|  |  | SC GA | $ 566722.58 | 16.4±7.0 | 9.8±4.4 | 1,240±15,596 | | |  |
|  |  | SC IFN β-1a | $ 924082.67 | 16.4±7.0 | 9.8±4.3 | 4,520±61,855 | | |  |
|  |  | IM IFN β-1a | $ 920472.98 | 16.4±7.0 | 9.8±4.4 | 4,527±61,854 | | |  |
|  |  | SC IFN β-1b | $ 855498.41 | 16.4±7.0 | 9.8±4.3 | 4,022±55,055 | | |  |
| Maciej J Maruszczak | QALY & ICER | Fingolimod | $  564448.36 | _ | 4.70 | 12,528 | | | Fingolimod remains cost-effective in highly active (HA) RRMS following the introduction of DMF to the UK market, and this model supports the evidence that has led it to be the only oral DMT reimbursed for HA RRMS in England. |
|  |  | dimethyl fumarate (DMF | $  549139.70 |  | 3.93 |  |  |  |  |
| Lorenzo Giovanni Mantovani | QALY, YLG ICER | Dimethyl fumarate | $  1396605.43 | 19.634 | 6.526 | Reference | | | This cost-effectiveness analysis confirms that dimethyl fumarate is an optimal first-line treatment for RRMS in Italy, compared with the other first-line alternatives. |
|  |  | IFN beta-1a – 22 mcg | $  1418953.20 | 19.533 | 5.786 | DMF dominates | | |  |
|  |  | IFN beta-1a – 44 mcg | $  1409201.85 | 19.600 | 6.189 | DMF dominates | | |  |
|  |  | IFN beta-1b – Betaferon | $  1474840.19 | 19.440 | 5.143 | DMF dominates | | |  |
|  |  | IFN beta-1b – Extavia | $ 1468349.53 | 19.440 | 5.143 | DMF dominates | | |  |
|  |  | Glatiramer acetate – 20 mg | $ 1454399.37 | 19.459 | 5.341 | DMF dominates | | |  |
|  |  | Teriflunomide – 14 mg | $ 1421793.87 | 19.547 | 5.953 | DMF dominates | | |  |
| Behzad Najafi | Health-related quality of life (HRQoL), ICER | CinnoVex | Annual per-patient cost: $2410 | _ | 69.5  for physical HRQoL &  63.3 for mental HRQoL | Reference | | | The results showed that CinnoVex was less expensive and more effective than Avonex over the study period. This implies that CinnoVex is a dominant option and there is no need to calculate the ICER. |
|  |  | Avonex | Annual per-patient cost: $4515 |  | 50.9  for physicalHRQoL &  56.6 for mental HRQoL | CinnoVex dominates | | |  |
| Mark J. C. Nuijten | QALY & ICER | preventive  treatment with interferon beta | $  455373.06 | _ | interferon group:  28.2 | $ 106076.04 per QALY | | | Preventive treatment with interferon beta in patients with multiple sclerosis may not be fully justified from a health-economic perspective, although interferon beta is associated with improved effectiveness compared with no preventive treatment. |
|  |  | No preventive treatment | $ 105319.26 |  | no-treatment group:  24.9 |  |  |  |  |
| Erkki Soini | QALY & ICER | DMF 240 mg PO BID | Total costs/patient, $  523140.50 | 12.098 | Total QALY/patient 7.808 | $ 51149.25 | $  114552.40 | | teriflunomide was less costly, with greater QALYs, versus glatiramer acetate and the IFNs. According to Bayesian treatment ranking (BTR), teriflunomide was the first-best among the disease-modifying therapies, with potential willingness-to-pay thresholds of up to €68,000/QALY gained. In the IIA (impact investment assessment), teriflunomide was associated with the longest incremental quality-adjusted survival and time without cane use. |
|  |  | teriflunomide 14 mg once daily | 512918.55 | 12.096 | 7.719 | $  36570.33 | vs. teriflunomide | |  |
|  |  | GA 20 mg SC once daily | 553208.02 | 12.087 | 7.475 | $  377612.44 | Dominant | |  |
|  |  | IFN-  β1a 44 mg SC TIW | 521832.96 | 12.092 | 7.595 | $  87610.24 | Dominant | |  |
|  |  | IFN-β1b 250 mg SC EOD | 613172.97 | 12.074 | 7.063 | Dom. | Dominant | |  |
|  |  | IFN-β1a 30 mg IM QW | 544899.55 | 12.088 | 7.456 | $  370707.19 | Dominant | |  |
|  |  | best supportive care (BSC)- placebo | 498725.36 | 12.084 | 7.331 | vs. BSC | $ 36570.33 | |  |
| Wenqing Su | QALY & ICER, HRQoL | DMF | $243,079 | 12.124 | 5.885 | Reference | | | DMF can be considered a  cost-effective option compared to other first-line DMTs |
|  |  | Glatiramer  Acetate (GA) | $219,741 | 12.105 | 5.357 | $44,118 | | |  |
|  |  | Rebif (Interferon-b 1a SC)  44 mcg | $240,134 | 12.116 | 5.610 | $10,672 | | |  |
| Xinke Zhang | QALY, ICER & incremental net  monetary benefit (INMB) | fingolimod | $239,947 |  | 3.69 | $  46,328 | | $  36,567 | Of the four DMDs, dimethyl fumarate is a dominant strategy to manage RRMS. Dimethyl fumarate dominated all other therapies over the range of willingness-to-pay (WTP) values. After dimethyl fumarate, teriflunomide was the most cost-effective therapy compared with IM IFN-b1a, with an incremental cost-effectiveness ratio of $7,115. |
|  |  | teriflunomide | $226,085 |  | 3.68 | $7,115 | | $  49,780 |  |
|  |  | dimethyl fumarate | $200,145 |  | 3.72 | Dominant | | $  80,611 |  |
|  |  | intramuscular (IM) interferon (IFN)-b1a | $223,606 |  | 3.34 | ICER vs. IM IFN-b1a | | INMB vs. IM IFN-b1a |  |
| Ray Gani  , et al | QALY & ICER | natalizumab |  |  |  | - | | | If UK society is willing to pay more than £8200 per QALY, or Health and Social Services are willing to pay more than £26 000 per QALY, this analysis suggests that natalizumab is likely to be a cost-effective treatment for all patients with RRMS |
|  |  | interferon-β |  |  |  | The ICER for natalizumab compared with interferon-β was £2300 per QALY. From a health and social care cost perspective, the ICERs were £18 700 per QALY | | |  |
|  |  | glatiramer acetate |  |  |  | The ICER for natalizumab compared with glatiramer acetate was £2000 per QALY. From a health and social care cost perspective, the ICERs were £20 400 per QALY | | |  |
|  |  | best supportive care |  |  |  | The ICER for natalizumab compared with best supportive care was £8200 per QALY. From a health and social care cost perspective, the ICERs were £25 500 per QALY, | | |  |
| Jim Chilcott, et all | cost per quality | Four disease-modifying treatments :  1-interferon betas 2-glatiramer acetate for relapsing-remitting and  3- interferon betas  4- glatiramer acetate for  secondary progressive multiple | 1-The base case cost per quality-adjusted life-year gained by using any of the four treatments ranged from £42 000 ($66 469; &61 630) to £98 000  based on efficacy information in the public domain 2-benefits increased the cost per QALY gained around 75%. 3- The estimates with a 20-year time horizon were markedly lower, ranging from £42 000 to £98 000 per QALY gained.  3- commercial in­confidence estimates of efficacy, the most favorable estimate is £35 000 per QALY and the least favorable is £104 000 per QALY |  |  | - | | | Cost-effectiveness varied markedly between the interventions. Uncertainty around point estimates was substantial., Price was the key modifiable determinant of the cost-effectiveness of these treatments |
| Julie Chevalier | QALY & ICER | DMF | $ 1191203.33 | - | 5.271 | - | | | Dimethyl fumarate can be considered a cost-effective option as it is on the efficiency frontier. |
|  |  | IFN beta-1a  44mcg | $  1185485.36 | - | 4.990 | - | | |  |
|  |  | IFNbeta-1a 30mcg | $  1191212.65 | - | 4.991 | - | | |  |
|  |  | IFN beta-1b, 250mcg | $  1207191.61 | - | 4.819 | - | | |  |
|  |  | Glatiramer  Acetate | $  1208023.54 | - | 4.950 | - | | |  |
|  |  | Teriflunomide | $  1192521.07 | - | 5.047 | - | | |  |
|  |  | Fingolimod | $  1267970.65 | - | 5.021 | - | | |  |
| Chalakorn Chanatittar  at, et al | ICER | BSC | BSC had the lowest cost = $235,000 |  | 49% | - | | | Compared with fingolimod and interferon treatments, BSC remains to be the most cost-effective treatment for RRMS in Thailand based on a WTP threshold of $4,500 per QALY gained. |
|  |  | fingolimod | the highest cost = $285,000 | 10.80 | 5.26  (%18) | $33,000 When compared with BSC | | |  |
|  |  | IFNβ−1b | - | - | %25 | $12,000 When compared with BSC | | |  |
|  |  | IFNβ−1a | - | - |  | $42,000 When compared with BSC | | |  |
| Murray G. Brown, et al | 1-Disability years avoided (DYA)  2- Cost per exacerbation avoided  3-ICER1 Foreign exchange rate used in the analysis was Can $1.00 D US $0.66 (1999). | interferon beta-1b | 1- Cost per disability year avoided before discounting is $124,892, and $181,395 after discounting at 5%  2- Total healthcare costs for all EDSS scores for Females Per person with MS: $1,976  Total healthcare costs for all EDSS scores for Males Per person with MS:$1,683 | - | - | - | | | Using the Expanded Disability Status Scale, cost per disability year avoided due to interferon beta-1b treatment in RRMS is quite high |
| Duygu Bozkaya, et al | ICER & EDSS = Expanded Disability Status Scale | natalizumab (NTZ) | Annual Drug cost: $71,773 | - | - | - | | | Costs ranged from $561,177 (NTZ) to $616,251 (GA). NTZ, DMF, and PEG were dominant (less costly and more effective) compared to FIN, GA, and IFN, respectively, for all ICERs. |
|  |  | fingolimod (FIN) | Annual Drug cost: $77922 | - | - | Incremental cost  NTZ vs FIN  -$35,524 | | |  |
|  |  | peginterferon beta-1a (PEG) | Annual Drug cost: $72,072 | - | - | - | | |  |
|  |  | subcutaneous interferon beta-1a (IFN, 44 mcg) | Annual Drug cost: $77797 | - | - | Incremental cost  PEG vs IFN-$37,790 | | |  |
|  |  | glatiramer acetate  (GA, 20 mg daily | Annual Drug cost: $80,436 | - | - | - | | |  |
|  |  | dimethyl fumarate (DMF) | Annual Drug cost: $73,371 |  |  | Incremental cost  DMF vs GA: -$47,573 | | |  |
| Mai F. Alsaqa’aby,  et al | ICERs and NMB. | Interferon 1a (Rebif 44 mcg) | $298 892 | - | 9.78 | - | | | 1-None of the DMDs were found to be cost-effective in the treatment of RRMS at a WTP threshold of  $100,000 in this analysis.  2- Monte Carlo simulation results showed that Rebif was the most cost-effective therapy at WTP of $50 000 with 95% probability.  3- Avonex reported the lowest ICER value of $337 282/QALY compared to Rebif as a common comparator.  4- The NMB of oral DMDs at a WTP of $100,000 (SAR375 000) was lower than the NMB of Rebif, showing that oral DMDs were a costly option.  would only be cost-effective at a WTP above  $300 000. |
|  |  | Teriflunomide | $360,631 | - | 9.72 | Dominated | | |  |
|  |  | Interferon 1a  (Avonex 30 mcg) | $374,502 | - | 10.01 | $337,282 | | |  |
|  |  | Fingolimod | $391,603 | - | 10.05 | $347,338 | | |  |
|  |  | Dimethyl Fumarate  (DMF) | $426,030 | - | 10.02 | $531,329 | | |  |
| Luis Hernandez,et al | QALY & ICER (All costs are  reported in 2014 US dollars) | peginterferon  beta-1a | - | - | Results Over 10 years, peginterferon beta-1a was dominant (i.e., more effective and less costly), with  cost-savings of $22,070 and an additional 0.06 QALYs when compared with interferon beta-1a 44mcg | - | | | This analysis suggests that long-term treatment with peginterferon beta-1a improves clinical outcomes at reduced costs compared with interferon beta-1a 44 mcg and glatiramer acetate 20mg and  should be a valuable addition to managed care formularies for treating patients with RRMS. |
|  |  | interferon beta-1a (44 mcg SC 3 times per week) | - | - | Results Over 10 years, | - | | |  |
|  |  | glatiramer acetate (20mg SC once daily) | - | - | peginterferon beta-1a was dominant (i.e., more effective and less costly), with  cost-savings  of $19,163 and 0.07, QALYs gained when compared with glatiramer acetate 20mg.- | - | | |  |
| Luis Hernandez, et al | QALY & ICER(All costs are reported in 2015 British Pounds) | Peginterferon beta-1a | Total cost : 106,843 | - | Total QALYs (patient-caregive): 7.32 |  | | | Long-term treatment with peginterferon beta-1a improves clinical outcomes, while its cost profile makes it either dominant or cost-effective compared with other self-injectable DMTs for the treatment of RRMS in Scotland. |
|  |  | Interferon beta-1a 30 mcg | Total cost: 113,257 | - | Total QALYs (patient-caregiver): 6.88 |  | | |  |
|  |  | Interferon beta-1a 22 mcg | Total cost: 115,614 | - | Total QALYs (patient-caregiver): 6.99 |  | | |  |
|  |  | Interferon beta-1a 44 mcg | Total cost: 112,523 | - | Total QALYs (patient-caregiver): 7.01 |  | | |  |
|  |  | Interferon beta-1b | Total cost: 110,657 | - | Total QALYs (patient-caregiver): 6.88 |  | | |  |
|  |  | Glatiramer acetate 20 mg | Total cost: 104,441 | - | Total QALYs (patient-caregiver): 6.90 |  | | |  |
| Aseel Bin Sawad, et al | QALY & ICER | Strategy 1: SM (symptom management) | US$161,136.60 | - | 10.49 | 2,297,141.53 comparing Strategy  2 to Strategy 1, | | | Strategy 1 was the cost-effective strategy for the treatment of relapsing-remitting multiple sclerosis when compared with other strategies. |
|  |  | Strategy 2: SM and  IFN-β-1a | US$551,650.66 | - | 10.66 |  |  |  |  |
|  |  | Strategy 3: SM and natalizumab | US$703,463.60 | - | 10.69 | -1,623,918.00 comparing Strategy 4 to Strategy 3. | | |  |
|  |  | Strategy 4: SM and alemtuzumab | US$670,985.24 | - | 10.71 |  |  |  |  |
| Hashemi-Meshkini A, et al | QALY (with currency  rate of 29,000 Iranian Rial (IRR) to 1 USD in 2016)& (ICER) | PEG-interferon : | 1- total discounted cost PEG-interferon: 68,688  USD  2-In each arm, cost of PEG-interferon 99% total cost | - | 5709.88 | 1- (ICER)  was estimated around 11111 US dollars (USD) per QALY gained for the  PEG-interferon vs. interferon  2- ICER (USD per QALY): cost discount rate (5%)= 12080  3- ICER (USD per QALY): Utility discount rate (3%)= 10208 | | | PEG interferon  beta 1 -a could be considered as a cost-effective treatment for Iranian patients with MS. |
|  |  | Interferon | 1-total discounted cost in interferon arm:  59,308 USD  2- In each arm, interferon beta 1a were  around and 97%total cost | - | 4865.61 |  |  |  |  |
