## Supplementary material for "Cost-Utility and Cost-Effectiveness Analysis of disease-modifying drugs of Relapsing-Remitting Multiple Sclerosis: A Systematic Review": Table 1

| **Topic** | **No.** | **Item** | **Reported?** |
| --- | --- | --- | --- |
| **TITLE** |  |  |  |
| **Title** | 1 | Identify the report as a systematic review. | Yes |
| **BACKGROUND** |  |  |  |
| **Objectives** | 2 | Provide an explicit statement of the main objective(s) or  question(s) the review addresses. | Yes |
| **METHODS** |  |  |  |
| **Eligibility criteria** | 3 | Specify the inclusion and exclusion criteria for the review. | Yes |
| **Information sources** | 4 | Specify the information sources (e.g. databases, registers) used to identify studies and the date when each was last searched. | Yes |
| **Risk of bias** | 5 | Specify the methods used to assess the risk of bias in the included studies. | No |
| **Synthesis of results** | 6 | Specify the methods used to present and synthesize results. | No |
| **RESULTS** |  |  |  |
| **Included studies** | 7 | Give the total number of included studies and participants and summarise relevant characteristics of studies. | Yes |
| **Synthesis of results** | 8 | Present results for main outcomes, preferably indicating the number of included studies and participants for each. If meta-analysis was done, report the summary estimate and confidence/credible interval. If comparing groups, indicate the direction of the effect (i.e. which group is favored). | No |
| **DISCUSSION** |  |  |  |
| **Limitations of evidence** | 9 | Provide a summary of the limitations of the evidence included in the review (e.g. study risk of bias, inconsistency, and imprecision). | No |
| **Interpretation** | 10 | Provide a general interpretation of the results and important implications. | Yes |
| **OTHER** |  |  |  |
| **Funding** | 11 | Specify the primary source of funding for the review. | Yes |
| **Registration** | 12 | Provide the registered name and registration number. | No |

Table 1. Evaluate this article based on the PRISMA checklist

**Table2.Quality Assessment**

| Item No | 1 | 2 | 3 | 4 | 5 | 6 | 7 | 8 | 9 | 10 | 11 | | 12 | 13a | 13b | 14 | 15 | 16 | 17 | 18 | 19 | 20 | | 21 | 22 | 23 | 24 | Total score | Overall quality |
| --- | --- | --- | --- | --- | --- | --- | --- | --- | --- | --- | --- | --- | --- | --- | --- | --- | --- | --- | --- | --- | --- | --- | --- | --- | --- | --- | --- | --- | --- |
|  |  |  |  |  |  |  |  |  |  |  | 11a | 11b |  |  |  |  |  |  |  |  |  |  |  |  |  |  |  |  |  |
|  |  |  |  |  |  |  |  |  |  |  |  |  |  |  |  |  |  |  |  |  |  | 20a | 20b |  |  |  |  |  |  |
| Reference |  | | | | | | | | | | | | | | | | | | | | | | | | | | | | |
| *1-Ray Gani*, et al ( 2008) | y | y | y | y | y | y | y | y | y | y | - | y | y | y | - | p | y | y | p | y | y | y | - | y | p | y | y | 22.5 |  |
| 2-Jim Chilcott, et all( 2003) | y | y | y | y | y | N | y | y | y | y | - | y | y | y | - | N | y | y | p | y | N | - | Y | y | N | Y | N | 18.5 |  |
| 3-Julie Chevalier(2016) | Y | Y | Y | Y | Y | Y | Y | Y | Y | Y | Y | - | Y | - | Y | Y | Y | Y | p | Y | Y | - | Y | - | P | N | N | 19.5 |  |
| 4-Chalakorn Chanatittarat, et al(2018) | Y | Y | Y | Y | Y | Y | Y | Y | Y | Y | Y | - | Y | - | Y | Y | Y | Y | P | Y | Y | - | Y | Y | N | P | N | 21 |  |
| 5-Murray G. Brown, et al (2000) | Y | Y | Y | Y | Y | Y | Y | Y | Y | Y | - | Y | N | - | Y | Y | Y | Y | P | Y | P | - | Y | Y | Y | Y | N | 21 |  |
| 6-Duygu Bozkaya,et al(2016) | Y | Y | Y | Y | Y | Y | Y | Y | Y | Y | N | N | N | - | Y | Y | Y | P | P | Y | Y | Y | - | Y | Y | Y | Y | 21 |  |
| 7-Mai F. Alsaqa’aby,et al (2017) | Y | Y | Y | Y | Y | Y | Y | Y | Y | Y | Y | - | Y | Y | - | Y | Y | Y | P | Y | Y | - | Y | Y | Y | Y | Y | 23.5 |  |
| Luis Hernandez,et al (2016) USA | y | y | y | y | y | y | y | y | y | y | - | y | y | y | - | y | y | y | p | y | y | - | y | y | y | y | N | 22.5 |  |
| Luis Hernandez, et al(2017) | y | y | y | y | y | y | y | y | y | y | y | - | y | y | - | y | y | y | p | y | y | y | - | y | y | y | N | 22.5 |  |
| 10-Aseel Bin Sawad, et al( 2017) | Y | Y | Y | Y | Y | Y | Y | Y | Y | Y | P | - | N | - | Y | Y | Y | P | P | Y | Y | - | Y | Y | Y | Y | N | 20 |  |
| 11-Hashemi-Meshkini A, et al (2018). | Y | Y | Y | Y | Y | Y | Y | Y | Y | Y | N | - | Y | Y | - | Y | Y | P | P | Y | Y | Y | - | Y | Y | Y | N | 21 |  |

Y= Yes, N=No, P = Partial, NA= Not applicable
