## Supplementary material for "Cost-Utility and Cost-Effectiveness Analysis of disease-modifying drugs of Relapsing-Remitting Multiple Sclerosis: A Systematic Review": Table 2

| **Table 2.** Study characteristics | | | | | | | | | | | |
| --- | --- | --- | --- | --- | --- | --- | --- | --- | --- | --- | --- |
| Reference  (year of publication) | Costing year | Setting | population | Compared interventions | Type of economic evaluation | Perspective | Time horizon | WTP  Threshold | Discount rate | Sensitivity analyses | Industry sponsorship |
| 1- Ray Gani, et al (2008) | 2005 | UK | 2048 MS patients | natalizumab compared with  interferon-β, glatiramer acetate and best supportive care | CEA (Markov model) | UK societal cost perspective | 30 years | £36,000 per QALY | 3.5% | Univariate sensitivity analysis | no support |
| 2- Jim Chilcott, et al (2003) |  | United  Kingdom | Participants Patients with relapsing-remitting multiple sclerosis and secondary progressive multiple sclerosis. | Four disease-modifying treatments: interferon betas, glatiramer acetate for relapsing-remitting and interferon betas, glatiramer acetate for secondary progressive multiple | CEA | UK NHS | 20 years | £20 000 | discounted costs at 6% per annum, the discounted quality of life benefits at 1.5% per annum | multivariate Monte Carlo sensitivity analysis | consultation with all stakeholders to its appraisals  process, using the best available evidence. |
| 3- Julie Chevalier (2016)  . | 2015 | French | 1,000 patients | glatiramer acetate, IFNbeta-1a 30mcg intramuscularly and 44mcg subcutaneously, IFN beta-1b 250mcg and teriflunomide as first-line therapies and fingolimod and natalizumab, as second-line therapies. | CEA | payer and societal | 30 years | - | 4% per annum during the first 30 years and2% after as requested by the French guidelines | Univariate and probabilistic | no support |
| 4- Chalakorn Chanatittarat, et al (2018) | 2011 TO 2014 | THAILAND | 105 MS patients aged 35 years | best supportive  care (BSC), fingolimod, IFNβ−1b, and IFNβ−1a | CUA (Markov model) | societal | month cycle length,  lifetime horizon or 30 years | WTP threshold of USD  4,500 per QALY gained. | 3 percent, Costs were converted to USD using 2016  average annual exchange rate of 35.26 Thai baht (THB) per 1  USD | univariate and probabilistic | no support |
| 5- Murray G. Brown, et al (2000) | 20 | Nova Scotia in Canada | 1,000 females  and 1,000 males followed 40 years | interferon beta-1b (IFN¯-1b) | CEA (simulation model) | Ministry of health  (MOH) | lifetime | - | 5% | One-way | no support |
| 6- Duygu Bozkaya, et al (2017) | 2016 | United States | for relapsing-remitting  multiple sclerosis (RRMS) | natalizumab (NTZ),  dimethyl fumarate (DMF), and peginterferon beta-1a (PEG) with fingolimod (FIN), glatiramer acetate  (GA, 20 mg daily), and subcutaneous interferon beta-1a (IFN, 44 mcg), | CEA (Markov Model) | third-party payer | Three-month cycles were modeled over a 10-year time horizon | - | 3% | One-way | no support |
| 7- Mai F. Alsaqa’aby, et al (2017) | 2015 | Saudi Arabia (Tertiary care hospital) | 1000 RRMS patients (for more than 400real MS patients) | oral agents v (fingolimod, teriflunomide, dimethyl fumarate,) vs. interferon (IFN)-b1 | CEA: Cohort Simulation Model (Markov Model( | Payer | 20 years and an annual cycle length. | $100,000 | 3%, All costs were  reported in Saudi Riyals (SAR) and converted into the equivalent value of 2015 US dollars | One-way and probabilistic (A probabilistic  sensitivity analysis based on a second-order Monte  Carlo simulation (1000 times)) | no support |
| 8- Luis Hernandez, et al (2016) | 2014 | United States | relapsing-remitting multiple sclerosis (RRMS) and includes adult patients. The population is 29.2% male with a mean age of 36.5 years | peginterferon beta-1a compared with interferon  beta-1a and glatiramer acetate | CEA( Markov cohort model) | US payer. | over 10 years | $50,000 | 3% | Probabilistic | no support |
| 9- Luis Hernandez, et al (2017) | 2015 | Scotland | relapsing-remitting multiple sclerosis (RRMS) | Peginterferon beta-1a and Interferon beta-1a 30 mcg and Interferon beta-1a 22 mcg and Interferon beta-1a 44 mcg and Interferon beta-1b and Glatiramer acetate 20 mg | CEA (Markov cohort model) | National Health Service and Personal Social Services | over 30 years | £20,000 per QALY | and discounted at 3.5% per year. | Probabilistic sensitivity analysis | Clemence Bougeard, Market Access  Analyst at Biogen, for her support obtaining the economic inputs for the  model |
| 10- Aseel Bin Sawad, et al (2017) | 2014 | USA | patients with RRMS Healthcare costs data were obtained from a study conducted in 2004 by Kobelt et al. assessing the cost of MS disease by stratified EDSS health states | Strategy 1: (symptom management [SM] alone), vs. Strategy 2: (SM and IFN-β-1a),  vs. Strategy 3 :(SM and natalizumab) vs. Strategy 4: (SM and alemtuzumab). | CEA (Markov model) | third-party payer | Over 20 years | $100,000 WTP threshold per QALY | 1- All costs were inflated to 2014 US$ by using the US\  2- costs were discounted using an annual discount rate of 3% | one-way, Probabilistic sensitivity analysis (second-order Monte Carlo simulation. | no support |
| 11- Hashemi-Meshkini A, et al (2018) | 2016 | Iran | 1,000 patients with relapsing-remitting MS (RRMS) | Pegylated versus non-pegylated interferon beta 1a | CEA (Markov model) | payer perspective (patients and third-party payers) | one-month cycles over 10 years | 15,945 USD | Cost discount rate (5%), Utility discount rate (3%) | One-way | Department of Pharmacoeconomics, Tehran University of Medical Sciences, Tehran, Iran, |
| Renée Else Michels, et al (2019) | 2016-2017 | Netherlands | Derived from a meta-analysis study (113 for cladribine group) | cladribine tablets vs. alemtuzumab and fingolimod | CEA & CUA | Societal | lifetime | €50,000/QALY gained | 4% for costs and 1.5% for outcomes | deterministic | Supported by Merck B. V group |
| Ali Imani, et al (2012) | 2011 | Iran | Model-based- population is not clear | Symptom Management vs. Avonex, Betaferon, Rebif, CinnoVex | CUA | Healthcare | lifetime | US$50,000/QALY gained | 7.2% annually | Performed | Tabriz University of Medical Sciences |
| Slobodan M. Janković, et al (2009) | 2009 | Serbia | Model-based - the population is not clear | Symptom management alone vs.  combination with subcutaneous glatiramer acetate (SC GA), subcutaneous interferon β-1a (SC IFNβ-1a), intramuscular interferon β-1a (IM IFNβ-1a), or subcutaneous interferon β-1b (SC IFNβ-1b). | CEA | Societal | lifetime (40 years) | WTP 5,000,000.00  RSD | 3% annually | multiple univariate sensitivity | Serbian Ministry of Science and Ecology |
| Maciej J Maruszczak, et al (2015) | 2013-2014 | England | Derived from a systematic review- the population is not clear | fingolimod vs. dimethyl fumarate (DMF) | CEA | NHS and Personal Social Services | Lifetime (50 years) | £20,000 and £30,000/QALY | 3.5% for both costs and benefits | Deterministic & Probabilistic sensitivity analysis | Novartis Pharmaceuticals UK Ltd, Camberley, UK |
| Lorenzo Giovanni Mantovani, et al (2019) | Euros inflated to June 2018 | Italy | Cohort and RCT based, the number of 1237 patients | Dimethyl fumarate vs. other first-line alternatives | CEA | Societal | Lifetime (50 years) | € 50,000 per QALY gained | 3.5% for both costs and outcomes | univariate deterministic and multivariate probabilistic | Biogen Italia (Milan, Italy) |
| Behzad Najaf, et al (2015) | 2012 | Iran | 140 patients | Avonex vs. CinnoVex | CEA | Ministry of Health and Medical Education | 1 year | Not clear enough | Not used | two-way sensitivity analysis | Iran University of Medical Sciences (IUMS) |
| Mark J. C. Nuijten, et al (2002) | 1998 | UK | The number of 560, 372, & 358 patients based on the previous three RCTs | preventive treatment with interferon beta,  No preventive treatment | CEA | third-party payer &  Societal | Lifetime (25 years) | Not stated | 6% annually | univariate sensitivity analyses | Not stated |
| Erkki Soini, et al (2017) | indexed to the 2014 price level | Finland | 713 patients | DMF 240 mg PO BID, teriflunomide 14 mg once daily,  GA 20 mg SC once daily, IFN-β1a 44 mg SC TIW,  IFN-β1b 250 mg SC EOD, IFN-β1a 30 mg IM QW, best supportive care (BSC)- placebo | CEA  /CBA | Finnish payer perspective. Scenario analysis with a societal perspective | 15 years | € 68,000 per QALY gained | 3% annually | Probabilistic sensitivity analysis | Sanofi Genzyme, Helsinki, Finland |
| Wenqing Su, et al. (2016) | 2013  Canadian dollars | Canada | Cohort and trial based 308 patients | DMF, Glatiramer Acetate (GA), Rebif (Interferon-b 1a SC) 44 mcg | CEA | Ministry of Health | Lifetime (20 years) | Canada ($50 000–60 000) | 5% for both health and economic outcomes | one-way and probabilistic sensitivity analyses | Supported by Biogen |
| Xinke Zhang, et al. (2014) | inflated to 2012  dollars | USA | A cohort of 1,000 patients | Fingolimod,  Teriflunomide,  dimethyl fumarate,  intramuscular (IM) interferon (IFN)-b1a | CBA/  CEA | societal | 5 years | US$ 150,000 per (QALY) | 3% annually | One-way  and probabilistic sensitivity analysis | No funding was received |
