## Supplementary material for "Cost-Utility and Cost-Effectiveness Analysis of disease-modifying drugs of Relapsing-Remitting Multiple Sclerosis: A Systematic Review": Figure 1

**Identification**

**Screeninging**

**Eligibilityity**

**Studies entered**

**Records identified through**

**database searching**

**‎ (n= 1360)‎**

**Records after duplicates**

**removed**

**‎(n = 891)‎**

**Number of studies to review the abstract**

**‎(n = 378)‎**

**Full text records included**

**(n= 115)‎**

**Records include in systematic review (n= 21)‎**

**‎ Number of studies excluded due to duplication**

**(n= 469)‎**

**Number of studies ‎removed due to unrelated ‎title‎**

**(n= 513)‎**

**Number of records ‎excluded due to unrelated ‎** **abstract ‎‎‎**

**(n= 263)‎**

**Number of records ‎excluded due to unrelated ‎** **Full text‎ ‎‎‎**

**(n= 94)‎**

**Figure 1.** PRISMA flowchart
